# Clemastine fumarate accelerates accumulation of disability in progressive multiple sclerosis by enhancing pyroptosis

**DOI:** 10.1101/2024.04.09.24305506

**Authors:** Joanna Kocot, Peter Kosa, Shinji Ashida, Nicolette Pirjanian, Raphaela Goldbach-Mansky, Karin Peterson, Valentina Fossati, Steven M. Holland, Bibiana Bielekova

## Abstract

Multiple sclerosis (MS) is an immune-mediated demyelinating disease of the central nervous system (CNS). Clemastine fumarate, the over-the-counter antihistamine and muscarinic receptor blocker, has remyelinating potential in MS. A clemastine arm was added to an ongoing platform clinical trial TRAP-MS (NCT03109288) to identify a cerebrospinal fluid (CSF) remyelination signature and to collect safety data on clemastine in patients progressing independently of relapse activity (PIRA). The clemastine arm was stopped per protocol-defined criteria when 3/9 patients triggered individual safety stopping criteria (χ^2^ p=0.00015 compared to remaining TRAP-MS treatments). Clemastine treated patients had significantly higher treatment-induced disability progression slopes compared to remaining TRAP-MS participants (p=0.0075). Quantification of ∼7000 proteins in CSF samples collected before and after clemastine treatment showed significant increase in purinergic/ATP signaling and pyroptosis cell death. Mechanistic studies showed that clemastine with sub-lytic doses of extracellular ATP activates inflammasome and induces pyroptotic cell death in macrophages. Clemastine with ATP also caused pyroptosis of induced pluripotent stem cell-derived human oligodendrocytes. Antagonist of the purinergic channel P2RX7 that is strongly expressed in oligodendrocytes and myeloid cells, blocked these toxic effects of clemastine. Finally, re-analyses of published snRNAseq studies revealed increased P2RX7 expression and pyroptosis transcriptional signature in microglia and oligodendrocytes in MS brain, especially in chronic active lesions. CSF proteomic pyroptosis score was increased in untreated MS patients, was higher in patients with progressive than relapsing-remitting disease and correlated significantly with rates of MS progression. Thus, pyroptosis is likely first well-characterized mechanism of CNS injury underlying PIRA even outside of clemastine toxicity.

**One Sentence Summary:** Clemastine enhanced disability accumulation in patients progressing by non-lesional MS activity by potentiating intrathecal P2RX7 signaling and pyroptosis.

## INTRODUCTION

Multiple sclerosis (MS) is chronic immune-mediated demyelinating disease of the central nervous system (CNS). Initial stages of MS are characterized by formation of focal inflammatory CNS lesions called MS plaques. Some MS plaques lead to subacute onset of neurological disability called MS relapse. All current disease-modifying treatments (DMTs) target this focal inflammatory “MS lesional activity” (i.e., MS plaques and relapses) very effectively; newer treatments inhibit formation of MS lesions by >95%. Despite this, the DMTs inhibit disability progression only modestly. In a meta-analysis of randomized clinical trials ^1^ DMT initiated after a mean age of 30 years do not inhibit disability progression completely; and their efficacy decreases linearly with the patient’s age. After mean age of 53 years, DMTs do not inhibit disability progression beyond placebo. Based on MS prevalence ^2,3^ this means that 45% of people living with MS today have no effective treatments and additional >40% of people with MS continue to accumulate disability on current DMTs.

Recognizing this unmet need, MS drug development focuses on targeting new disease mechanisms. Among these is remyelination, as chronically demyelinated axons are considered less resilient to neurodegenerative mechanisms compared to their myelinated counterparts ^4^. Clemastine fumarate, an over-the-counter antihistamine with anticholinergic properties against muscarinic M1 receptor, induces remyelination in-vitro ^5^, in multiple animal models ^6–9^ and in the randomized, blinded cross-over Phase II MS trial (ReBUILD; ClinicalTrials.gov ID NCT02040298 ^10^. In 50 MS patients with mean age of 40.1 years who had mild disability (i.e., mean Expanded Disability Status Scale [EDSS] 2.2 on ordinal scale from 0 to 10), clemastine demonstrated remyelinating effect. The primary outcome was drug-induced acceleration of the electrical conduction across visual pathways measured by the change in latency of the visual evoked potential (VEP) positive 100 msec wave (P100). MS patients with abnormally prolonged P100 (mean P100 = 128.6 and 126.8 msec in two cross-over groups) were enrolled. Clemastine-treated patients decreased P100 latency by 1.7 msec per eye (93 eyes total; p=0.0048). Interpreting measured mean P100 velocities as representing 28.6 to 26.8 msec delay due to MS, clemastine corrected on average 5.9-6.3% of this pre-existing abnormality. This was associated with a trend for low contrast visual acuity improvement by the mean of 0.9 letters/eye (p=0.085). Clemastine also improved brain myelin water fraction measured as reduction in radial diffusivity from diffusion tensor magnetic resonance imaging (MRI), but did not affect another MRI biomarker of demyelination, magnetization transfer ratios ^11^. No significant changes in clinical outcomes were observed in this short trial. None of the clinical trials testing other remyelinating agents showed clear efficacy ^12^.

The vulnerability of demyelinated axons to degeneration is not the only candidate mechanism of PIRA. Analyses of MS CNS tissue ^13,14^ and cerebrospinal fluid (CSF) biomarkers from MS patients identified compartmentalized inflammation in CNS tissue ^15,16^ insufficiently targeted by current DMTs ^17^ and varied neurodegenerative processes ^18–20^. These candidate mechanisms evolve with MS duration ^21^ and, analogous to the heterogeneity of the formation of acute MS lesions ^22^, are not uniformly detected in all MS patients. This observed heterogeneity of MS disease mechanisms implies that similarly to other chronic diseases with complex pathophysiology such as cardiovascular diseases, patient-specific “custom combination treatments” may be required to inhibit disability progression in MS. The prerequisite for development of such personalized treatments is the ability to measure candidate disease mechanisms comprehensively, in living people. To achieve this goal in 2017 we initiated a platform, open-label, CSF biomarker-guided clinical trial: Targeting Residual Activity by Precision, biomarker-guided combination therapies of Multiple Sclerosis (TRAP-MS; ClinicalTrials.gov ID NCT03109288). The objectives of TRAP-MS are: 1. To develop clinical trial methodology that allows economical screening of prospective therapeutic agents for their efficacy on biological processes related to MS severity using CSF biomarkers; 2. To develop knowledge base of intrathecal effects of current DMTs and novel treatments targeting varied mechanisms of MS progression; and 3. To establish and validate framework for development of effective combination therapies for MS.

Based on the results of ReBUILD trial, in 2020 we added a clemastine arm to the TRAP-MS trial with the goal of identifying CSF remyelination signature and extending the safety/tolerability data on the long-term use of clemastine in patients progressing by PIRA (i.e., by “non-lesional” MS activity). We stopped the clemastine arm in 2022 due to protocol-defined safety criteria. This paper identifies clemastine-enhanced pyroptosis as mechanism for observed toxicity. Most importantly, we show that pyroptosis associated with smoldering intrathecal inflammation is likely mechanism of PIRA in general.

## RESULTS

### Clemastine arm of TRAP-MS protocol was stopped after three out of nine patients triggered safety stopping criterion

TRAP-MS patients are monitored with clinical and imaging safety outcomes every 6 months and with CSF biomarkers 6 months after each therapeutic change. Clinical safety is monitored by a continuous, machine-learning derived Combinatorial Weight Adjusted Disability Scale (CombiWISE; range 0-100). CombiWISE correlates highly with EDSS (R^2^=0.93, p<0.0001; Fig. 1A) and reproducibly measures yearly disability progression in small (∼30 subjects) MS cohorts^23^. To receive treatment under TRAP-MS protocol, patients on stable DMT (or untreated if desired) must be progressing by at least 0.5 CombiWISE units/year, derived from the patient-specific linear regression models based on a minimum of 4 clinic visits with each visit ≥6 months apart (i.e., minimum of 18 months; Fig. 1B). This represents “baseline” progression slope, whereas analogously derived data from on-treatment visits measures “therapy” progression slopes.

**Fig. 1.**
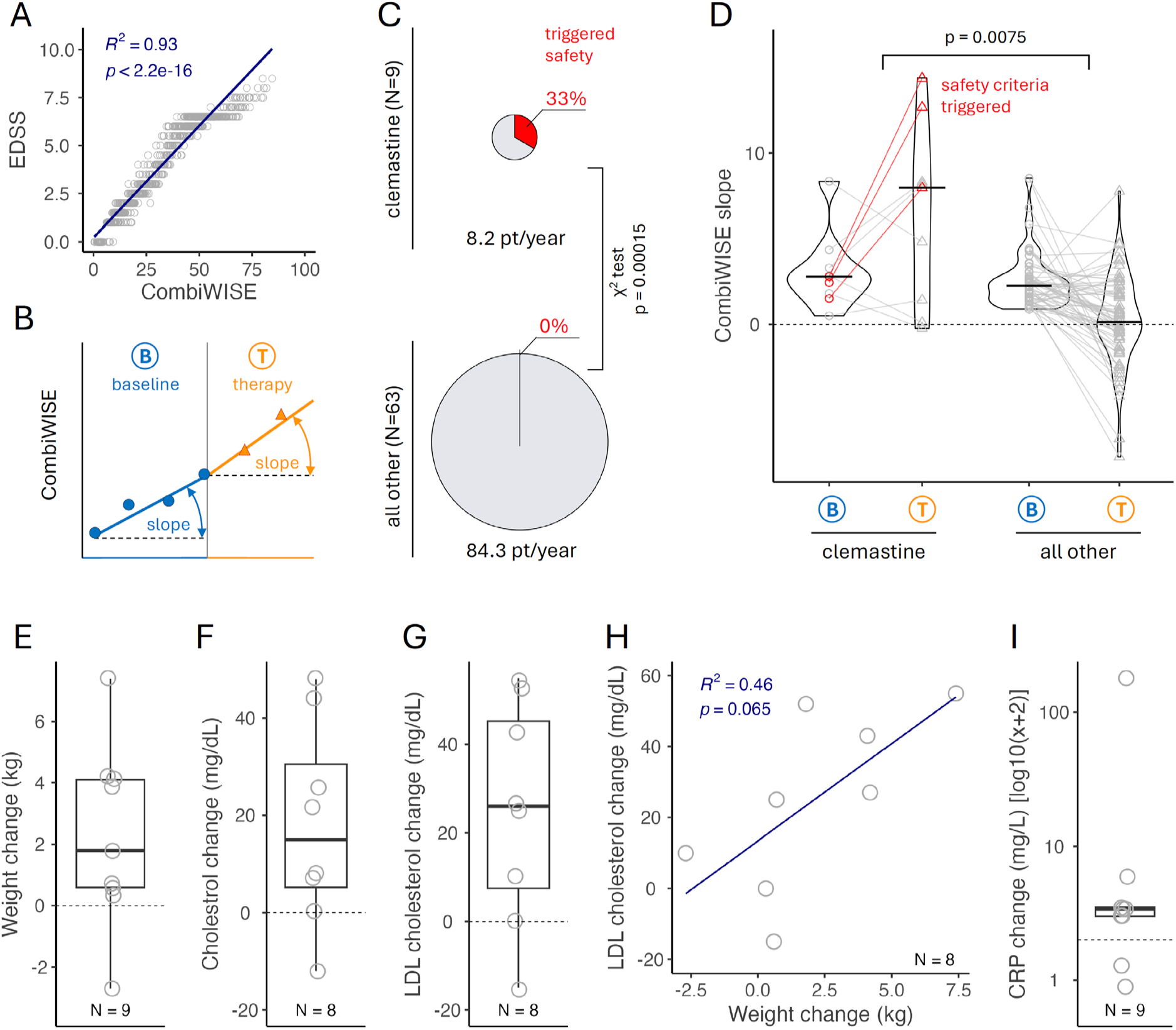
Clemastine-induced changes in disability, metabolism, and inflammatory markers. (**A**) Clinical safety was monitored by a continuous, machine-learning derived CombiWISE scale (range 0-100) that correlates strongly with EDSS (range 0-10). (**B**) A minimum of 4 visits on stable therapy spanning at least 18 months is required to measure baseline (blue) CombiWISE slope. On-therapy CombiWISE slope (orange) is calculated based on 6-months follow-up data collected after theory initiation. (**C**) 3 out of 9 patients on clemastine therapy triggered safety criteria, in contrast to none of the 63 patient-specific treatments of 6 other TRAP-MS therapies; p-value of probability of this occurrence was based on χ^2^ test. (**D**) Progression CombiWISE slopes at baseline (B) and therapy (T) were compared between clemastine arm and all other 6 TRAP-MS therapies. Black horizontal line represents median value of the group. Red color indicates 3 patients that triggered safety stopping criteria treatment slope exceeding 5x baseline slope. Displayed p-value was generated from two-sided unaired Wilcoxon rank test comparing therapy-induced change in CombiWISE slope between the clemastine arm and all other therapies. Clemastine induced increase of weight (**E**), total cholesterol (**F**), and LDL cholesterol (**G**) levels between baseline and treatment and these changes showed strong association as seen on example of weight change versus LDL cholesterol change (**H**). Furthemore, clemastine induced increase of inflammatory biomarker CRP (**I**). Lipid panel was an optional laboratory test and the results are missing for one patient. EDSS – Expanded Disability Status Scale, CombiWISE – Combinatorial Weigh-adjusted Disability Scale, LDL – low density lipoprotein, CRP – C-reactive protein, Blue line in H represents linear regression line, gray dashed line in D-G and I represents 0 change. The lower and upper hinges of the boxplots correspond to the first and third quartiles (the 25th and 75th percentiles). The upper and lower whiskers extend from the hinge to the largest and smallest value, respectively.

TRAP-MS protocol has pre-defined individual and treatment arm safety stopping criteria. Individual patients must stop the study drug when on-treatment yearly CombiWISE progression slope exceeds 5x the baseline progression slope, or when on-treatment accumulation of new/enlarging MS lesions exceeds 3x the baseline average. Individual stopping criteria were based on natural history cohort simulations from 536 patient/years, where they occur only in 3.4% of all clinic visits. Treatment arm is stopped for toxicity when 3 treated patients trigger individual stopping criterion.

Sixteen patients initiated clemastine treatment, with nine completing at least one 6-months follow-up visit, (total of 8.2 patient/years) when 3/9 (33.3%) fulfilled individual stopping criteria of on-treatment progression slopes exceeding baseline progression slopes by more than 5-fold (Fig 1C). The probability of seeing this result by chance is 0.015% (p=0.00015; χ^2^ test comparing clemastine arm with remaining TRAP-MS treatments, where 0 out of 63 treatment-specific patient slopes (representing 42 patients) triggered safety criteria after 84.3 patient/years on remaining 6 tested drugs: montelukast, losartan, hydroxychloroquine, pioglitazone, dantrolene and pirfenidone). This triggered per-protocol closing of the clemastine arm for toxicity (table S1).

Apart from the incidence of patients triggering individual stopping criteria, Fig 1D shows that most (6/9) clemastine-treated patients progressed at higher than baseline rates, while most remaining TRAP-MS patients slowed rates of disability progression (p=0.0075 comparing change in slopes for clemastine-treated versus remaining patients. Note that TRAP-MS trial is ongoing, and no treatment arm reached pre-defined numbers to analyze clinical outcomes. Therefore, we can’t exclude that the apparent benefit of remaining drugs is due to placebo effect universally observed in MS trials).

Based on very low probability that observed results are due to chance, together with vastly different treatment effects on progression slopes between clemastine and other TRAP-MS treatments, we conclude that clemastine accelerated disability progression in these patients with advanced progressive MS.

### Clemastine-treated patients developed metabolic syndrome with activation of innate immunity

The patients who reached stopping criteria tended to be older (median age 71.4 vs 60.6 years), heavier (median weight 93.8 vs 75.5 kg), and more disabled (i.e., median EDSS 7.0 vs 6.5 and median CombiWISE 60.9 vs 52.9) compared to subjects who did not. However, the adverse effects of clemastine were seen in most treated patients: we observed an increase in the CombiWISE slope in 78%, weight gain in 89%, increased total cholesterol in 88% and low-density lipoprotein (LDL) cholesterol in 88% of clemastine-treated patients (Fig. 1E-G). These metabolic changes were correlated: e.g., an increase in serum LDL cholesterol explained 46% of variance of weight gain (Fig. 1H). Furthermore, 78% of clemastine-treated patients increased serum C-reactive protein (CRP) (Fig. 1I). These changes were not observed on other TRAP-MS tested drugs.

To summarize, clemastine-treated patients exhibited systemic signs of metabolic syndrome and acute phase response suggestive of innate immunity activation.

### Analysis of CSF biomarkers shows that clemastine treatment altered purinergic metabolism

To understand intrathecal effects of clemastine, we measured relative concentrations of 7,000 CSF proteins from CSF samples collected before and after clemastine therapy using a multiplex DNA-aptamer (SOMAscan) assay.

We scaled the measured CSF biomarkers to HD values (to standardize them for direct comparison), and then calculated change in biomarker levels between therapy and baseline CSF samples in 9 patients that received clemastine therapy. We did not expect any significant differences for individual proteins after adjustment for multiple comparisons, because the number of measured proteins greatly exceeds the number of patients/samples, and indeed, none of the clemastine-induced changes passed the FDR-adjusted threshold of p-value < 0.05 (table S2).

However, gProfiler analysis of clemastine-induced changes in biomarkers with the greatest effect size (those exceeded two standard deviations above and below the mean, representing 5% of biomarkers outside of the normal distribution) unexpectedly identified several significant changes induced by clemastine in GO terms related to purine metabolism: e.g., purine nucleotide binding (GO:0017076; p= 3.43xe-3), purine nucleotide metabolic process (GO:0006163; p=1.83e-4) and purine-containing compound biosynthetic process (GO:0072522; p=1.00e-3).

We conclude that clemastine alters metabolism of purine ribonucleotides.

### Clemastine potentiates ATP binding to purinergic P2RX7 receptor and induces activation of inflammasome and pyroptosis in macrophages

While presenting our unexpected results from clemastine arm of TRAP-MS trial at an internal research meeting, one of the coauthors (S.H.) alerted us to the fact that clemastine enhances killing of intracellular pathogens such as mycobacteria in myeloid cells based on its potentiating effect on ATP-signaling via purinergic P2RX7 receptor ^24,25^. While clemastine does not activate P2RX7 by itself, it acts as allosteric modulator, opening the channel in response to suboptimal/non-activating concentrations of extracellular ATP.

In immune cells, P2RX7 is highly expressed in myeloid lineage, including microglia (https://gtexportal.org/home/gene/P2RX7; https://www.proteinatlas.org/ENSG00000089041-P2RX7). P2RX7 activates myeloid cells in response to danger-associated molecular pattern (DAMP) of high extracellular ATP, usually caused by lytic (immunogenic) cell death such as necrosis, necroptosis or pyroptosis. P2RX7 signaling mediates efflux of intracellular K^+^, activating the NOD-like receptor pyrin domain-containing protein 3 (NLRP3) inflammasome. Assembled inflammasome activates caspase-1 leading to extracellular release of active IL-1β and IL-18, master-regulators of the innate immunity. Inflammasome activation requires two signals: 1. Priming by pathogen-associated molecular patterns (PAMPs; e.g., lipopolysaccharide [LPS]) or proinflammatory cytokines. Priming initiates transcription of inflammatory mediators, including NLRP3 components and pro-IL-1β. 2. Activation (e.g., by DAMPs such as ATP) leads to inflammasome assembly resulting in proteolytic cleavage of pro-caspase 1. Active caspase 1 cleaves other substrates such as gasdermin D (GSDMD), enabling formation of mitochondrial and plasma membrane channels, through which active IL-1β and other inflammatory stimuli are released. Prolonged P2RX7 signaling leads to pyroptotic cell death, further exacerbating inflammation by releasing active caspase 1 and DAMPs such as ATP and nuclear high mobility group box 1 (HMGB1) protein ^26,27^.

With this background, we asked whether clemastine activates inflammasomes (measured by release of IL-1β) in LPS-primed myeloid cells activated by suboptimal ATP concentrations. In the human monocytic leukemia cell line THP-1, used for inflammasome/pyroptosis studies ^28^, clemastine increased release of IL-1β and active caspase-1 (compared to ATP+DMSO control) already 3h post-activation (Fig. 2A and 2C, respectively). These effects were mediated by GSDMD channel, as GSDMD deficient THP-1 cells released significantly less active caspase-1 (Fig. 2D and fig. S1A) and IL-1β (Fig. 2B). Lack of GSDMD significantly attenuated but did not abolish the potentiating effect of clemastine on IL-1β secretion, suggesting that other channels, e.g., GSDME, may be involved. Clemastine also enhanced immunogenic cells death of THP-1 cells (reflected by the loss of cell membrane integrity measured by cellular uptake of SytoxGreen (Fig. 2I), release of lactate dehydrogenase (LDH; Fig. 2E) and decreased cell viability (Fig. 2G), again in GSDMD-dependent manner (Fig. 2F and 2H), proving pyroptotic cell death.

**Fig. 2.**
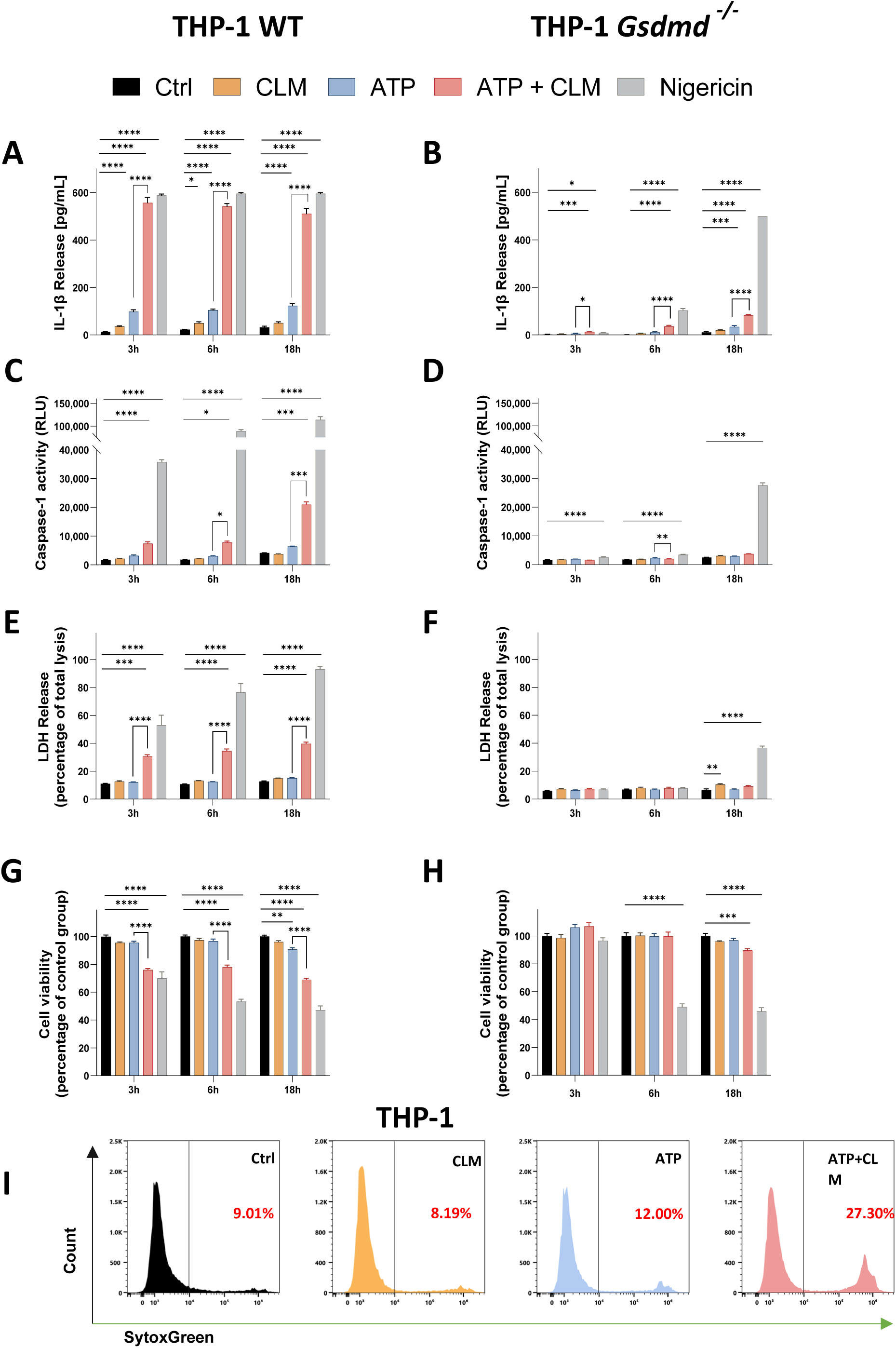
Clemastine (CLM) potentiation effect on ATP-induced inflammasome activation and GSDMD-driven pyroptotic cell death in THP-1 cells. The THP-1 cells were primed with 200 ng/mL LPS overnight and then treated either with medium + DMSO (negative control, Ctrl), 10 µg/mL CLM, 2 mM ATP +/-10 µg/mL CLM or 2.5 µM Nigericin (positive control) for 3h, 6h, and 18 h. Release of the pro-inflammatory cytokine IL-1β into the culture medium of THP-1 cells (**A**) and THP-1 *Gsdmd ^-/-^* cells (**B**), determined by ELISA assay. Activity of Caspase-1 in the culture medium of THP-1 cells (**C**) and THP-1 *Gsdmd ^-/-^* cells (**D**), determined using bioluminescent assay. Lytic cell death of THP-1 cells (**E**) and THP-1 *Gsdmd ^-/-^* cells (**F**), determined by LDH activity released into the culture medium. Cell viability of THP-1 cells (**G**) and THP-1 *Gsdmd ^-/-^* cells (**H**) evaluated by the MTS assay. Data are presented as mean ± SEM of three independent experiments, each performed in duplicate (n=6). One way ANOVA followed by Dunnett’s multiple comparisons test was used to compare the testing groups with control group (Ctrl). One-way ANOVA test followed by Holm-Sidak’s multiple comparison was used to compare ATP and ATP+CLM group (to assess the potentiation effect of CLM). *, P ≤ 0.05. **, P ≤ 0.01. ***, P ≤ 0.001. ****, P ≤ 0.0001. (I) Flow cytometry analysis of cell uptake of SYTOX green in THP-1 cells primed with 200 ng/mL LPS overnight and then treated either with medium + DMSO (control), 10 µg/mL CLM or 2 mM ATP +/-10 µg/mL for 90 min. Representative plot of two independent experiments.

Next, we confirmed these observations in primary human macrophages. Under identical conditions, in contrast to THP-1 cells, ATP alone caused lytic cell death in monocyte-derived macrophages, but the effect was further potentiated by clemastine in time-dependent manner (Fig. 3A to E). Compared to ATP alone, clemastine significantly enhanced pyroptosis of macrophages assessed by an increased release of both active caspase-1 (Fig. 3B and fig. S1B) and LDH into culture supernatants (Fig. 3C), increased cell membrane permeability (Fig. 3E) and a decreased cell viability (Fig. 3D).

**Fig. 3.**
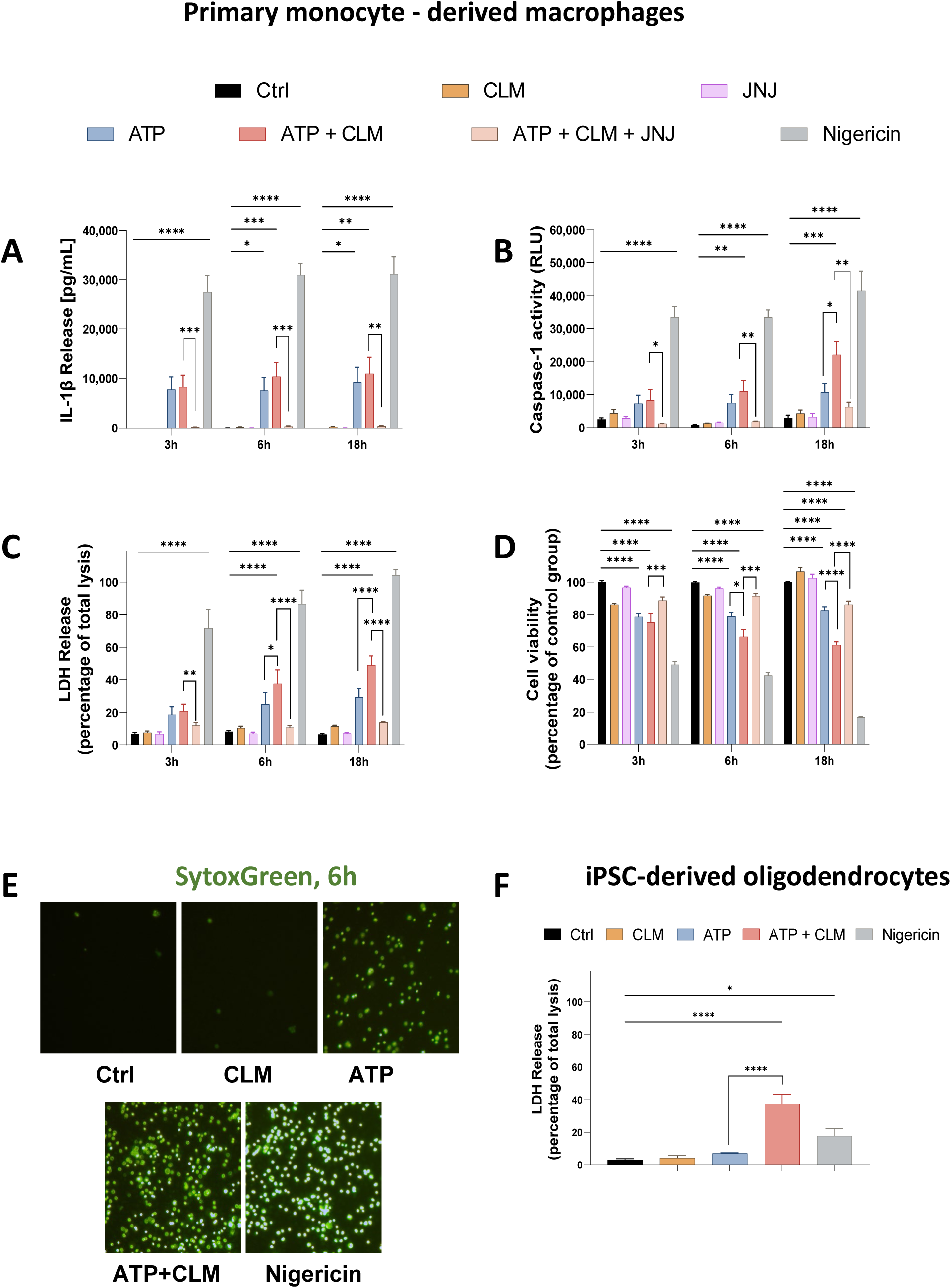
The potentiation effect of Clemastine (CLM) on ATP-induced lytic cell death in primary monocyte-derived macrophages and iPSC-derived oligodendrocytes. The primary monocyte-derived macrophages (**A-D**) were primed with 200 ng/mL LPS overnight, then pre-treated with 30 nM JNJ-54175446 (a selective purine P2X7 receptor antagonist) for 1 h, followed by treatment either with medium + DMSO (negative control, Ctrl), 10 µg/mL CLM, 2 mM ATP +/-10 µg/mL CLM or 10 µM Nigericin (positive control) for 3h, 6h, and 18 h. Levels of the pro-inflammatory cytokine IL-1β (**A**) and Caspase-1 (**B**) activity in the culture medium determined by ELISA and bioluminescence assay, respectively. Lytic cell death (**C**) determined by LDH activity released into the medium and cell viability (**D**) evaluated by the MTS assay. Data are presented as mean ± SEM of three independent experiments, each performed in duplicate (n=6). Representative fluorescence microscopy images depicting SYTOX Green-stained MDMs treated with drugs for 6 h (**E**). iPSC-derived oligodendrocytes (**F**) were primed with 200 ng/mL LPS overnight followed by treatment either with medium + DMSO (negative control, Ctrl), 10 µg/mL CLM, 2mM ATP +/-10 µg/mL CLM or 10 µM Nigericin (positive control). LDH release after the treatments for 18 h (**F**). Data are presented as mean ± SEM of four independent samples. One way ANOVA followed by Dunnett’s multiple comparisons test was used to compare the testing groups with control group (Ctrl). One-way ANOVA test followed by Holm-Sidak’s multiple comparison was used to compare ATP and ATP+CLM group (to assess the potentiation effect of CLM). *, P ≤ 0.05. **, P ≤ 0.01. ***, P ≤ 0.001. ****, P ≤ 0.0001.

In contrast to THP-1 cells, primary human macrophages released higher quantities of IL-1β in response to ATP alone and clemastine potentiated IL-1β release only marginally (Fig. 3A and fig. S2).

To confirm that the potentiating effects of clemastine on pyroptosis of primary macrophages were P2RX7 dependent we pretreated macrophages with a selective brain-penetrant P2RX7 antagonist JNJ-54175446 (Fig. 3A to D) and observed significant attenuation of lytic cell death.

To further confirm our results, we performed the bulk RNA-seq analysis of primary human macrophages to identify signaling pathways affected by clemastine (+ATP) stimulation. GSEA revealed that ATP alone induces upregulation of mitochondrial dysfunction and apoptosis pathways (fig. S3) but inflammasome activation, pyroptosis (fig. S4 and S5) and immunogenic cell death (fig. S6) were only seen in ATP+clemastine condition.

We conclude that in the proinflammatory environment with extracellular ATP, clemastine significantly enhances inflammasome activation and pyroptotic cell death of myeloid cells.

### Clemastine potentiates immunogenic cell death of human oligodendrocytes

Intriguingly, oligodendrocytes express the highest P2RX7 mRNA levels among human cell types and the spinal cord has the highest P2RX7 tissue expression (https://gtexportal.org/home/gene/P2RX7; https://www.proteinatlas.org/ENSG00000089041-P2RX7). Because the clemastine-induced disability progression was clinically localized to the spinal cord, we hypothesized that oligodendrocytes are susceptible to pyroptosis in a proinflammatory environment with high extracellular ATP concentrations.

Thus, we differentiated human oligodendrocytes from induced pluripotent stem cells (iPSCs) using a published protocol ^29^ and tested whether oligodendrocytes are susceptible to lytic cell death in the presence of extracellular ATP. We found that while insufficient alone, 2 mM ATP caused the lytic cell death of oligodendrocytes in the presence of clemastine (Fig. 3F).

We conclude that in a pro-inflammatory environment with sub-lethal concentrations of extracellular ATP, clemastine causes lytic cell death in oligodendrocytes.

While pyroptotic cell death of oligodendrocytes is the most plausible mechanism of the observed clemastine toxicity, it remains unclear whether pyroptosis contributes to progression of MS disability in patients not treated with clemastine.

### P2RX7 expression and pyroptosis signaling pathway are increased in microglia and oligodendrocytes in MS brain tissue

To investigate whether P2RX7 expression associates with transcriptional pyroptosis signature in MS CNS, we re-analyzed publicly available single-nucleus RNA sequencing (snRNAseq) datasets.

We found enhanced P2RX7 expression in MS compared to control brains (Fig. 4A). Chronic active MS lesions, followed by periplaque white matter and chronic inactive lesions had higher P2RX7 expression than active lesions. We confirmed high expression of P2RX7 in oligodendrocytes. Within the oligodendrocyte subtypes, P2RX7 was most highly expressed in the OLG6 subtype which we found highly enriched in MS tissue (Fig. 4B).

**Fig. 4.**
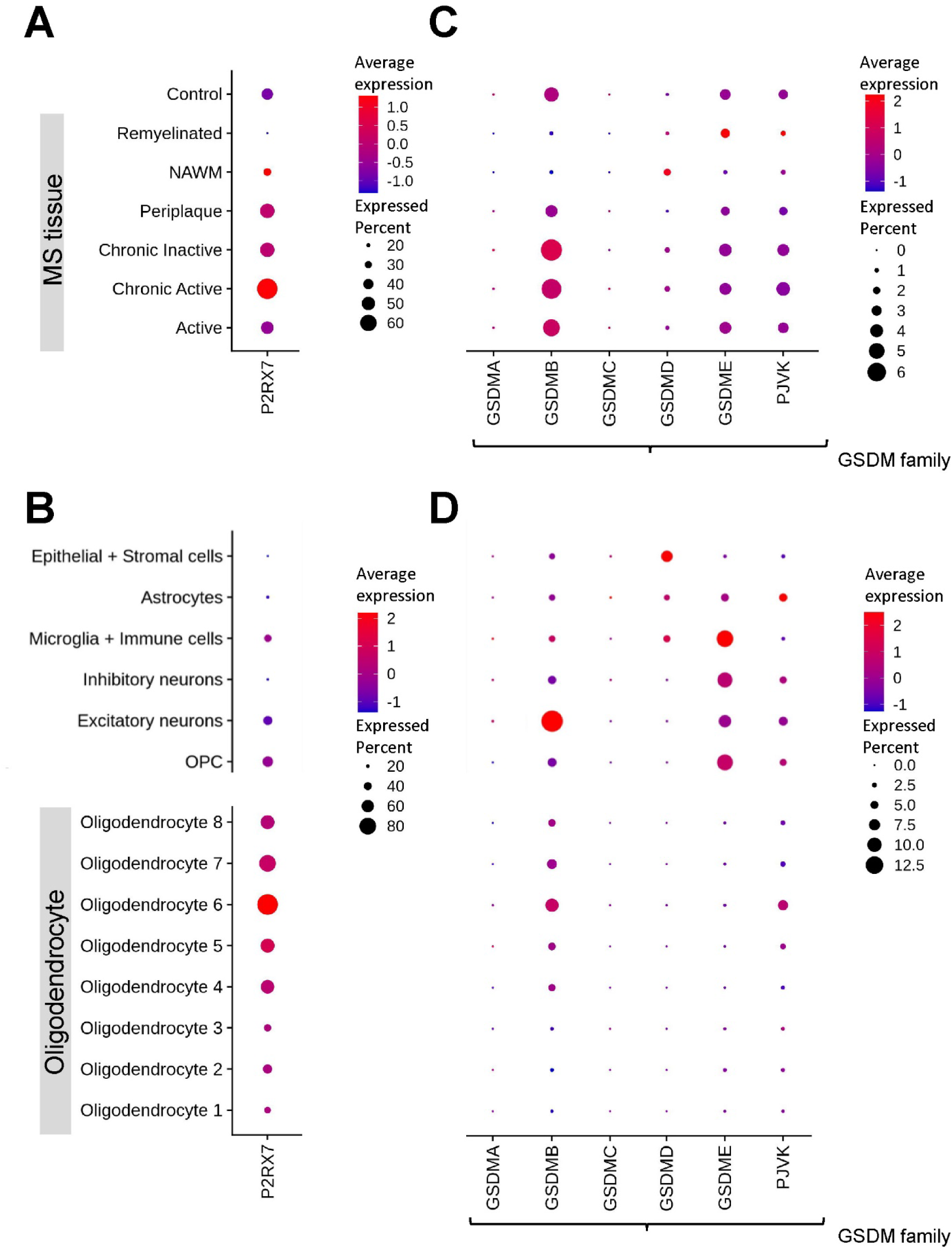
Gene expression associated with pyroptosis signaling pathway in CNS tissue. Panels (**A)** and (**C)** illustrate the profiles of P2RX7 and GSDM family across the lesion types. Meanwhile, panels (**B)** and (**D)** show gene expressions based on cell types. P2RX7: purinergic receptor P2X7; GSDM: gasdermin; PJVK: pejvakin; NAWM: normal-appearing white matter; OPC: oligodendrocyte precursor cells.

Assessing the spectrum of gasdermin channels we found GSDMB most prevalent, especially in chronic active, chronic inactive and active MS lesions (Fig. 4C). Consistent with public databases, oligodendrocytes expressed GSDMB and Pevjakin (PJVK) gasdermin channels. We found the highest expression of these in the aforementioned OLG6 subgroup, greatly expanded in MS compared to non-MS CNS (Fig. 4D). On the other hand, GSDMD was expressed in microglia and epithelial/stromal cells and GSDME in microglia, neurons, and astrocytes, but at levels comparable between MS and control tissues (fig. S7).

We integrated transcriptomic data for all known proteins involved in pyroptosis into a pyroptosis transcriptomic activation score (see Methods). The pyroptosis transcriptomics score was highest in microglia/macrophages (12.42 in MS vs 8.6 controls; 44% increase in MS), then epithelial and stromal cells (8.1 MS vs 7.88 controls) and finally in oligodendrocytes, where the pyroptosis score was 173% higher in MS compared to non-MS cells (4.69 MS vs 1.72 controls).

We conclude that MS microglia/macrophages and particular subgroup of oligodendrocytes greatly expanded in MS have higher transcriptomic signatures of pyroptosis in comparison to control cells. Genes associated with pyroptosis show the highest transcription levels in chronic active MS lesions, followed by periplaque white matter and chronic inactive lesions.

### Pyroptosis signaling pathway measured by CSF proteins positively correlates with MS severity. Compared to placebo, clemastine therapy significantly activates intrathecal pyroptosis

To assess whether pyroptosis contributes to progression of neurological disability in non-clemastine treated MS patients, we calculated a proteomic pyroptosis score (see Methods for detail) from CSF proteomic data measured by DNA-aptamers (i.e., SOMAscan) in a cross-sectional cohort of 168 MS patients, in a cohort of 72 longitudinally followed MS patients (assigned to placebo arms of two double-blind clinical trials of progressive MS: NCT00950248 ^30^ and NCT01212094 ^17^), and 49 healthy donors (HD) (for demographic data see Table S1).

Consistent with our re-analysis of CNS tissue transcriptomics data, we observed a significant increase in the proteomic pyroptosis scores in MS versus HD CSF (p=1.1×10^-15^, Fig. 5A). Moreover, the CSF pyroptosis score was higher in progressive MS than in relapsing-remitting multiple Sclerosis (RRMS) patients (Fig. 5B, median 2.32 vs 1.23, p=1.9×10^-5^) and was elevated in baseline (pre-treatment) CSF samples in all three patients in TRAP-MS clemastine arm that triggered safety stopping criteria (Fig. 5C). When comparing yearly change in CSF pyroptosis score calculated from longitudinal CSF in MS patients assigned to the placebo arm of previous progressive MS trials with clemastine arm of TRAP-MS, we observed significant increase in CSF pyroptosis signature in clemastine-treated patients (Fig. 5D, median 0.03 vs 0.41, p=0.032).

**Fig. 5.**
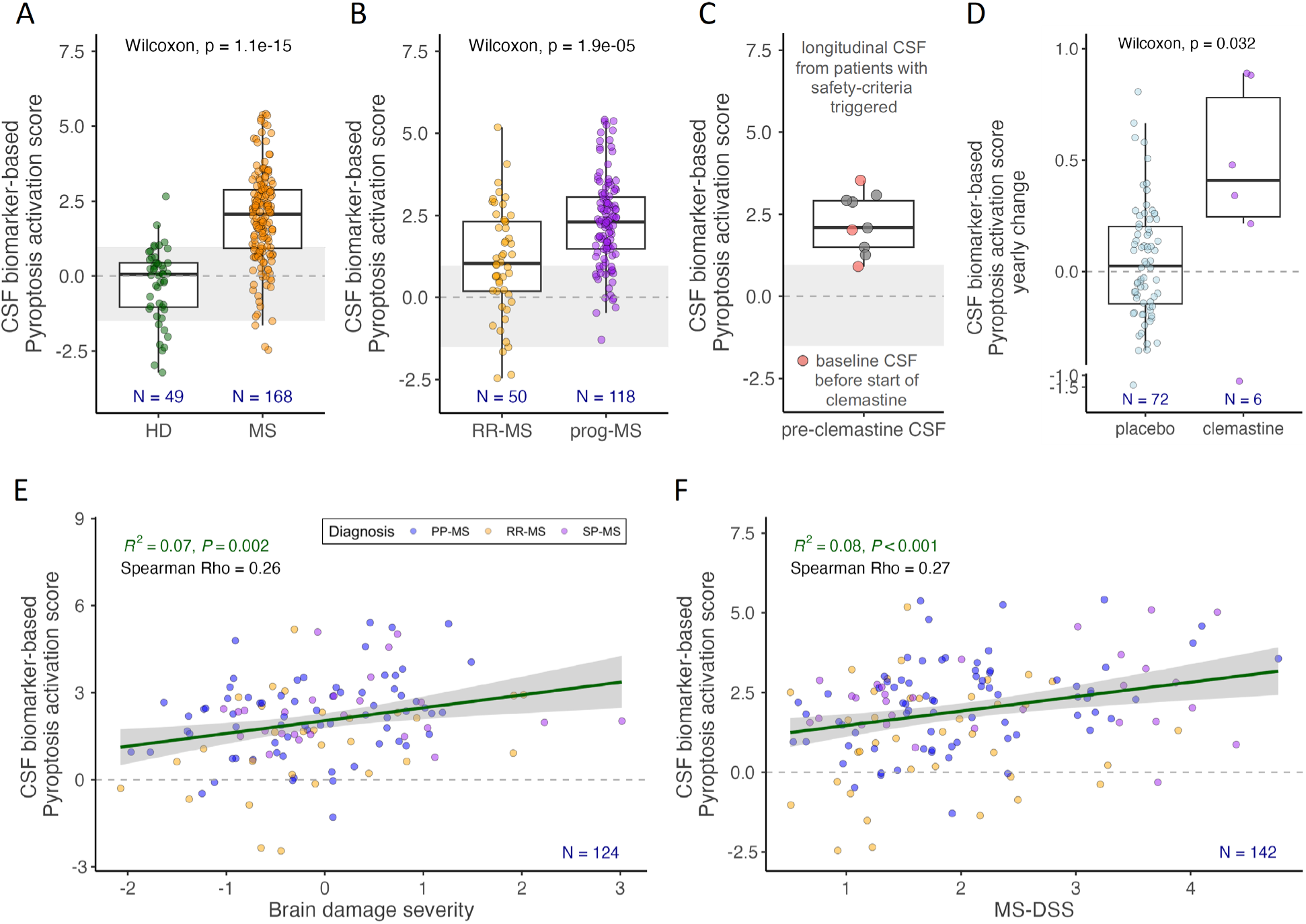
MS severity associated Pyroptosis signature in CSF increases with Clemastine treatment. CSF biomarker-derived Pyroptosis activation score is significantly elevated in MS patients compared to HD (**A**) and in progressive MS (prog-MS) patients compared to relapsing-remitting MS (RR-MS) patients (**B**). The gray area represents HD mean +/-standard deviation. (**C**) Longitudinal pre-clemastine CSF samples of patients that triggered safety criteria show elevated Pyroptosis activation score. CSF samples collected just prior starting clemastine are highlighted in red. (**D**) Clemastine-induced yearly change in Pyroptosis activation score is significantly elevated compared to untreated MS patients. The lower and upper hinges of the boxplots correspond to the first and third quartiles (the 25th and 75th percentiles). The upper and lower whiskers extend from the hinge to the largest and smallest value, respectively. Pyroptosis activation scores is strongly associated with the rate of accumulation of cognitive and physical disability, represented by Brain damage severity (**E**) and MS-DSS (**F**), respectively. The green line depicts linear regression line, and the gray shaded area shows 95% confidence interval. The gray dashed line depicts 0.

Thus, we conclude that compared to healthy volunteers, MS patients have CSF-proteomic signature indicative of intrathecal pyroptosis, that this signature is significantly higher in patients with progressive MS compared with RRMS and that clemastine further increases the intrathecal proteomic pyroptosis signature in vivo.

Furthermore, in the longitudinal MS cohort the CSF pyroptosis activation score correlated significantly (Fig. 5E; Spearman Rho = 0.26, R^2^ = 0.07, p=0.002) with rates of brain tissue loss measured as Brain damage severity score derived from volumetric MRIs (see Methods).

Analogously, the CSF pyroptosis activation score also correlated with the MS disease severity score (MS-DSS; Fig. 5F, Spearman Rho = 0.27, R^2^ = 0.08, p<0.001), which integrates clinical and imaging data, reflects past and predicts future rates of disability accumulation in the independent validation cohort ^31^.

Positive correlation of the CSF pyroptosis activation score with MS severity outcomes together with other presented results implicate pyroptosis is important pathogenic mechanisms of PIRA-associated CNS tissue destruction in MS.

## DISCUSSION

In this study we report that clemastine increases the rate of disability accumulation in patients with advanced MS progressing by PIRA, by facilitating prolonged opening of P2RX7 channel, potentiating intrathecal pyroptosis of myeloid cells and oligodendrocytes in MS CNS. In science, we quantify the probability of obtaining observed results by chance using p-values. While in isolation every piece of data we present maybe considered improbable but perhaps still possible by chance, collectively our data fit a unified story we find impossible to dismiss. Considering two obvious limitations of our study, that we 1. Do not have CNS tissue from clemastine-treated subject to demonstrate drug-induced oligodendroglial cell death and 2. Did not run clinical trial of brain-penetrant P2RX7 inhibitor, which is the only way one could prove pathogenicity of P2RX7-mediated pyroptosis in MS, data we did collect provide robust practically obtainable evidence supporting our conclusions.

The strong counterargument is that clemastine toxicity was not identified in the ReBUILD trial that included more MS patients. Indeed, we found this observation so reassuring that we attributed the unusual rates of disability progression in the first two safety criteria-triggering clemastine arm patients to the weight gain from a sedentary lifestyle during the COVID19 pandemic. But we knew we carefully selected safety criteria based on internal natural history data to uncover drug toxicity on MS progression with high sensitivity and accuracy. Furthermore, equally disabled patients treated in parallel with alternative drugs in TRAP-MS platform trial did not experience analogous disability worsening. Therefore, after the third patient triggered safety criterion, we stopped clemastine arm for toxicity in accordance with approved protocol and focused on gaining mechanistic understanding of this toxicity.

Designing TRAP-MS trial with the belief that patients’ safety always takes precedence over scientific needs, we were doubtful that resulting limited number of clemastine treatment CSF samples would elucidate toxicity mechanism(s). Instead, every data we obtained, both in vivo and in-vitro, fitted like puzzle pieces into unified story. After stopping the clemastine arm, the first unexplained observations we presented to the Data and Safety Monitoring board were those of metabolic syndrome observed in clemastine-treated subjects. Next, unbiased CSF proteomics showed enhanced purinergic signaling, observed only in clemastine-treated patients. While our infectious disease colleagues were aware of the potentiating effect of clemastine on P2RX7 ATP-binding ^24,25^ because they used clemastine to enhance mycobacteria killing in human macrophages, none of the publications describing remyelinating effects of clemastine identified or discussed this off-target effect, greatly relevant to MS. Likely nobody in MS field, us included, knew about this potential clemastine toxicity until we collected data presented here.

By not activating P2RX7 directly, clemastine may be toxic only in subjects with high CNS extracellular ATP levels. The rapid flow of vascular extravasate observed as dynamic contrast-enhancement in acute MS lesions likely precludes focal accumulation of extracellular ATP necessary to sustain P2RX7 signaling. Whereas, in the smoldering inflammation associated with chronic active MS lesions or periplaque white matter, such focally high ATP levels may occur when oligodendrocytes are in vicinity of activated myeloid cells and extracellular fluid flows slowly. This conclusion is in line with the first-in-human P2RX7 positron emission tomography (PET) tracer study in 5 RRMS patients and 5 healthy controls that observed decreased binding of [^11^C]SMW139 in contrast enhancing lesions, but increased binding in MS white matter ^32^.

This explains why we observed higher transcriptomics pyroptosis score in chronic active/inactive MS lesions and periplaque white matter when compared to active MS lesions and why CSF proteomic pyroptosis score is also significantly higher in progressive MS than RRMS. This also explains why much older (mean age of 62.5 vs 40.1 years) and more disabled (i.e., mean baseline EDSS 5.7 vs 2.2) TRAP-MS patients were more susceptible to the toxic effect of clemastine compared to RRMS patients in ReBUILD trial. Clemastine is highly lipophilic ^25^, which may increase its concentration in CNS white matter and in fat tissues, where it may aggravate metabolic syndrome. Our data suggest that pre-existing obesity may be another susceptibility factor for clemastine toxicity, perhaps by increasing inflammasome activation of adipose tissue macrophages and monocytes, which may travel to CNS. The other differences between ReBUILD and TRAP-MS trials were treatment duration (2-3 months vs >6 months) and more granular clinical scale to identify safety signal in TRAP-MS (i.e., change in EDSS versus change in CombiWISE slopes). Indeed, we show in the fig. S8A-C that using only change in EDSS would not identify clemastine toxicity.

Considering that clemastine fumarate is an over-the-counter antihistamine sometimes used off-label based on promising ReBUILD trial, our unfortunate experience highlights potential for off-target effects of (most) small molecules. One cannot assume that a drug is safe for new indication because it is available over the counter. Likewise, safety cannot be extrapolated from studies that enrolled different patient populations, including different disease stages.

The silver lining in this story is that clemastine toxicity helped to identify intrathecal P2RX7-linked pyroptosis as a likely PIRA mechanism even outside of clemastine treatment. Although linking CSF pyroptosis activation score to CNS pathology in living people is not possible, our data strongly suggest that MS patients with CSF proteomic pyroptosis signature are enriched for subjects progressing by PIRA and losing brain volume (i.e., Brain damage severity score) without forming new MS lesions. In fact, most patients in TRAP-MS trial do not form new MS lesions despite measurable disability progression on CombiWISE. The formation of new MS lesions triggers escalation of DMTs, because current DMTs suppress MS lesional activity at all MS stages ^33^. Thus, TRAP-MS patients are progressing by “non-lesional” MS activity (which encompasses PIRA), either because the MS lesion formation is fully suppressed by DMTs or because patients with advanced age stopped forming new lesions. Our retrospective application of CSF proteomic Pyroptosis score showed that 83% of these PIRA-experiencing patients who eventually received clemastine (including 3 patients who triggered safety criteria) had elevated CSF pyroptosis score *before* initiating clemastine treatment.

Location of the brain transcriptional pyroptosis signature indicates that this lytic cell death occurs in MS tissue where oligodendrocytes are in vicinity of smoldering inflammation characterized by activated microglia and macrophages. These myeloid cells undergo vicious circle of inflammasome activation in response to inflammatory stimuli up to the point of succumbing to pyroptosis and releasing active caspase-1, IL-1β, IL-18 and DMAPs such as extracellular ATP and nuclear antigen HMGB1. Neighboring oligodendrocytes, having unusually high P2RX7 expression, open P2RX7 channels in response to extracellular ATP, releasing intracellular K^+^ and dying by pyroptosis if P2RX7 channel opening is prolonged, further fueling (sterile) inflammation. Oligodendrocytes may be uniquely susceptible to P2RX7-mediated pyroptosis not only because of their high P2RX7 expression, but also because in contrast to most human cells, oligodendrocytes lack ATP-degrading ecto-nucleotidases NT5E (CD73; https://www.proteinatlas.org/ENSG00000135318-NT5E) and ENTPD1 (CD39; https://www.proteinatlas.org/ENSG00000138185-ENTPD1).

Although earlier pathology studies identified inflammasome activation and pyroptotic cell death in microglia ^34^ and oligodendrocytes ^35^ in MS CNS, current study makes four critical contributions: 1. It proposes that inhibiting P2RX7-mediated pyroptosis of oligodendrocytes is the desired effect of P2RX7 blockage in MS (as opposed to inhibiting inflammasome activation of microglial/macrophages); 2. It links intrathecal pyroptosis in living subjects to clinical and imaging outcomes of PIRA and MS severity; 3. It proposes that P2RX7 inhibitors may have stronger therapeutic effect in progressive, compared to RRMS patients; 4. It develops CSF pyroptosis score that can identify patients with therapeutic target and that can be used as pharmacodynamic (PD) marker in Phase II human trials.

Our conclusion that P2RX7-mediated pyroptosis in oligodendrocytes is one of the candidate mechanisms of PIRA is supported by informative cuprizone demyelination study, where P2RX7-deficient mice were robustly protected against toxic demyelination. On the other hand, density of myeloid cells in cuprizone demyelinated tissue was only marginally lower in P2RX7-deficient animals. Gene expression profiling of microglia purified from demyelinated tissue showed that pro-inflammatory phenotype of myeloid cells, including their expression of inflammasome-related genes, was not affected by the lack of P2RX7 ^36^. In view of our results, we reinterpret the results of this cuprizone study as supportive of the primary pathogenic role of P2RX7 signaling on oligodendrocytes, not on microglia. Indeed, direct triggering of P2RX7 in isolated rat optic nerve caused MS-like focal demyelinated lesions with associated axonal injury ^37^. Nevertheless, only oligodendrocyte-specific P2RX7 deficiency could prove primary pathogenic role of excessive P2RX7 signaling in in-vivo demyelination.

An important question that neither our, nor these animal studies addressed, is what physiological role P2RX7 plays in oligodendrocytes? Because P2RX7 expression is part of the Myelin sheath organization cluster (https://www.proteinatlas.org/ENSG00000089041-P2RX7), long-term P2RX7 blockage may negatively alter myelin biology. Indeed, in rats, P2RX7 protein is present both in oligodendrocytes and compact myelin (visualized by electron microscope immunogold staining) ^37^. P2RX7 is also expressed in myelinated Schwan cells ^38^, where it affects developmental myelination of peripheral nerves. Reassuringly, blocking P2RX7 during remyelination phase of aforementioned cuprizone study did not alter remyelination and people with hypomorphic P2RX7 alleles do not have clinically relevant myelin pathology (see below).

The pathogenic role of P2RX7 in MS progression (and neurodegeneration in general) is also supported by P2RX7 genetic polymorphism ^39–41^, where gain-of-function variants are observed at higher frequencies, and hypomorphic variants at lower frequencies in different neurodegenerative diseases but also in faster aging ^42^. Nevertheless, the largest MS genetic study did not link P2RX7 polymorphism to MS severity ^43^. The heterogeneity and multiplicity of candidate pathogenic mechanisms ^4,21,44,45^ and their evolution during MS natural history, compounded by inadequate accuracy of traditional MS severity outcomes make studies of MS severity extremely challenging. Thus, this issue needs more research.

Proving pathogenicity of intrathecal pyroptosis outside of clemastine toxicity requires interventional clinical trial that successfully blocks pyroptosis in MS CNS. Pharmaceutical industry developed several P2RX7 inhibitors, some brain-penetrant (e.g., JNJ-54175446, GSK314181A, AZ11645373, AZD9056 and CE-224535 ^46^). P2RX7 inhibitors were tested in systemic autoimmune/inflammatory diseases such as rheumatoid arthritis and Crohn’s disease. Based on clinicaltrials.gov, it seems that most P2RX7 inhibitors were abandoned after disappointing Phase II trials ^47,48^ or PD modeling ^49^. To our knowledge, only brain-penetrant JNJ-54175446, used in our in-vitro experiments, is currently in clinical development for depression ^50^. The negative P2RX7 inhibitor trials are concerning, but they targeted NLRP3 inflammasome, which have multiple/redundant activation modes. In contrast, we show that MS patients have increased intrathecal pyroptosis, which (for CSF proteomic pyroptosis score) correlates with rates of MS progression and is transcriptionally linked to P2RX7 expression in subpopulation of oligodendrocytes greatly expanded in MS CNS. Furthermore, the high P2RX7 expression together with the lack of ecto-nucleases CD39 and CD73 make oligodendrocytes uniquely vulnerable to P2RX7-mediated death. We hope our results will lead to proof-of-principle clinical trial of brain-penetrant P2RX7 inhibitor in MS. Pre-selecting patients using CSF proteomic pyroptosis score and measuring drug-induced change in this CSF signature as PD marker for rapid yes-no decisions would de-risk more expensive trials, necessary to measure the effect of P2RX7 inhibition on PIRA.

## MATERIALS AND METHODS

### Study design

This study aimed to identify CSF remyelination signature and collect the safety data on the long-term use of clemastine in MS patients progressing by PIRA. Details of the trial design are described in fig. S9 and the Subjects and Samples processing paragraphs below. For in-vitro experiments, three independent experiments were conducted with two technical replicates to demonstrate biological reproducibility and ensure adequate statistical power for comparisons. The exception was the experiment for iPSC-derived oligodendrocytes, performed once with four technical replicates. The figure legends indicate the group, sample sizes, and statistical tests used. No outliers or other data points were excluded.

### Subjects

MS patients and HD were prospectively recruited between January 2008 and January 2023 as part of the natural history protocol “Comprehensive Multimodal Analysis of Neuroimmunological Diseases of the Central Nervous System” (Clinicaltrials.gov identifier NCT00794352). For details see demographic data in Table S1. Informed consent was obtained from all patients. All subjects underwent a comprehensive clinical, MRI, laboratory evaluation, and research lumbar puncture at the protocol entry, with optional follow-up visits. This protocol also served as a screening platform for identification of MS patients who would fulfill inclusion criteria for enrollment into the protocol “Targeting Residual Activity By Precision, Biomarker-Guided Combination Therapies of Multiple Sclerosis (TRAP-MS, Clinicaltrials.gov identifier NCT03109288, for protocol design, see fig. S9). Sixteen MS patients, fulfilling the inclusion criteria, were enrolled into the clemastine arm of the TRAP-MS protocol. Nine of those completed at least one follow-up visit scheduled every 6 months. 42 patients were enrolled into additional six treatment arms (dantrolene, hydroxychloroquine, losartan, montelukast, pioglitazone, and pirfenidone) of TRAP-MS. CSF was collected at the baseline (within one year before treatment initiation) and 6 months after treatment start.

### Clinical outcomes

Neurological exams were performed by MS-trained clinician and recorded into NeurEx^TM^ App ^51^ that automatically generates MS disability scores, such as EDSS. CombiWISE ^23^, Brain Damage severity ^44^, and MS Disease Severity Score (MS-DSS) ^31^ were calculated as described. Briefly, CombiWISE is a machine-learning derived progression outcome that combines disability levels measured by EDSS, Scripps Neurological Rating Scale ^52^, timed 25 foot walk and non-dominant hand nine hole peg test. Brain Damage Severity is calculated as age-residuals of Brain Damage disability outcome that combines progression measured by cognitive test Symbol Digit Modalities Test (SDMT) and MRI volumetric outcome Brain Parenchymal fraction (proportion of brain tissue volume to the whole brain volume). MS-DSS is a machine learning-derived severity outcome that combines disability level measured by CombiWISE, age, therapy history, and levels of CNS tissue destruction measured by MRI.

### Sample processing

CSF was collected on ice and processed immediately according to a written standard operating procedure by investigators blinded to diagnoses, clinical, and imaging outcomes. Aliquots were assigned alphanumeric identifiers and centrifuged for 10 minutes at 300xg at 4°C within 30 minutes of collection. The cell-free supernatants were aliquoted and stored in polypropylene tubes at −80°C.

### SOMAScan analysis

CSF protein content of MS patients and HD was analyzed by SOMAScan® technology (SomaLogic®, Boulder, CO) using the version 4.1 of the assay that measures relative fluorescent units (RFUs) of 7,550 epitopes. The RFUs have been mathematically processed to normalize and calibrate the signal within and between different plates using control samples embedded in each 96 well plates.

### Inflammasome activation – in-vitro mechanistic studies

THP-1 and THP1-KO-GSDMD cells were seeded in 96-well plates at 3×10^5^/well and primary MDMs at 1×10^5^/well and left for 4 h. Then, for inflammasome activation, the cells were primed with 200 ng/mL LPS overnight, followed by activation with 10 µg/mL clemastine (CLM), 2 mM ATP +/-10 µg/mL CLM or Nigericin (as positive control) for 3h, 6 h or 18 h. We used 2.5 µM Nigericin for THP-1 and THP1-KO-GSDMD cells and 10 µM Nigericin for primary MDM cells and iPSC-derived oligodendrocytes. The cells primed with LPS without any activation treatment served as negative control (Ctrl). JNJ-54175446, a selective purine P2X7 receptor (P2X7R) antagonist at 30 nM, was used in the experiments involving primary MDM cells.

Human iPSC-derived oligodendrocytes were generated using previously published protocol ^29^. On day 70 of differentiation, iPSC-derived oligodendrocytes were treated as above for 18h. After drug incubation, the conditioned medium was collected, centrifuged at 250 x g to remove cell debris, and either used immediately or frozen at −80°C until analysis. Inflammasome activation was evaluated by levels of the pro-inflammatory cytokine IL-1β and the activity of caspase-1 released to the cell culture medium, while pyroptotic cell death was assessed by Sytox Green uptake, LDH release, or cell viability. Details on the in-vitro experiments are reported in the Supplementary material.

### Statistical analysis

#### Identification of Clemastine-induced changes in CSF proteome

The HD-scaled levels of Somamers were compared before and 6 months after clemastine treatment and Somamers changes outside of 95% of the normal distribution (mean +/-2*standard deviation) were evaluated by g:Profiler version e111_eg58_p18_30541362 to identify gene ontology terms and pathways that were significantly changed by clemastine treatment. Clemastine-induces changes in Somamer levels were also tested by one sample Wilcoxon test, with a null hypothesis of 0 change in somamer values between baseline and therapy sample. Both raw p-values and FDR-adjusted p-values were recorded.

#### CSF biomarker-based Pyroptosis activation score calculation

To calculate Pyroptosis activation score in CSF, first, we intersected known genes participating in pyroptosis signaling based on the knowledge base of Ingenuity Pathway Analysis (IPA®) platform with CSF proteins measured by the SOMAScan assay. Out of those, 10 proteins demonstrated statistically significant correlation with global MS severity outcome (sum of scaled values of Brain damage severity ^44^, CombiWISE severity ^44^, and MS-DSS ^31^) and were selected for the Pyroptosis activation score calculation (Table S3). The CSF Pyroptosis activation score was then calculated as a sum of HD-scaled Somamer levels of the selected 10 proteins, multiplied by the correlation coefficient of each protein with the global MS severity outcome.

#### In-vitro mechanistic experiments

Statistical analysis of data from in-vitro mechanistic studies was performed using GraphPad Prism 9 (GraphPad Software, Inc.). Data are presented as means ± SEM. One-way analysis of variance (ANOVA) followed by either Dunnett’s test (to compare various treatment groups with the control group) or Holm-Sidak’s test (to compare two experimental groups) was used, as indicated in figure legends. P < 0.05 was considered statistically significant.

#### Bulk RNA-seq analysis

Paired end FASTQ files were generated and concatenated by cat package. Preprocessing for FASTQ files (quality control, adapter trimming, quality filtering, per-read quality pruning of data) was performed by fastp package ^53^. The expression of transcripts was quantified based on GRCh38.p14 by using Salmon package ^54^. Preprocess for FASTQ files and quantifying of gene expression were performed by cloud based Galaxy (v0.23.2) ^55^. Raw reads count data was normalized and comparison was performed by DESeq2 in iDEP 0.96 ^56^. K-means clustering, principal component analysis was done in iDEP. Significant gene expression change was defined as log2FoldChange >0.6 and False discovery rate (FDR) < 0.05. Data were analyzed using the QIAGEN’s Ingenuity Pathway Analysis suite (IPA, QIAGEN, www.qiagen.com/ingenuity). For details on sample preparation, RNA extraction, library preparation and sequencing, see methods in the supplementary material.

#### Reanalysis of single-nucleus RNA sequencing

##### Published data collection

Published data of single-nucleus RNA sequencing (snRNA-seq) from MS and control (GSE180759, GSE118257, and PRJNA544731) was re-analyzed, consisting of total of 21 MS subject and 17 controls (Age: 46.7 ± 6.9 vs. 55 ± 14.5 years old, female proportion 10/21 (48%) vs. 6/17 (35%), respectively). Mean MS disease duration is 19 ± 8.8 years and included 2 PPMS and 19 SPMS patients.

##### Clustering and annotation of nucleus

Each raw data was counted, and QC was checked by Cellranger ver.7.0 (10x Genomics). Count data reads were merged, and principal component analysis (PCA) and t-distributed stochastic neighbor embedding (tSNE) were analyzed by Seurat (ver. 4.2.0) ^57^. The nuclei with high mitochondrial gene content (>10%) or high unique molecular identifier (UMIs) (>10,000) were excluded, leaving 361,253 nuclei for downstream analyses. Each dataset was merged using Canonical correlation analysis (Seurat, RPCA method). The merged dataset was clustered by Seurat, K-nearest neighbor (KNN) graph and tSNE. Non-biased clustering was done by weighted nearest neighbor analysis, and each cluster was annotated based on variable gene expression. The nucleus was classified as oligodendrocyte progenitor cell (OPC), oligodendrocyte (OLG), excitatory neurons, inhibitory neurons, microglia/immune cells, astrocytes, and epithelial/stromal cells. OLG were clustered into 8 subtypes (OLG1-8). Each gene expression was assessed based on 1: Subject type (MS vs. control), 2: Lesion type (active lesion, chronic active lesion, chronic inactive lesion, control lesion, NAWM: normal appearing white matter lesion, peri plaque lesion, remyelinated lesion), and 3: Clustered cell type.

### Transcriptional Pyroptosis pathway activation (tPPAS) score calculation in snRNA-seq

To assess cellular contribution to the pyroptosis pathway, we calculated tPPAS for each gene expression type. First, we downloaded all genes included in the Pyroptosis pathway from IPA® software, together with their directionality (i.e., transcription level correlating positively or negatively with pathway activation). For genes that lacked this directionality information we used directionality of correlation coefficient of analogous CSF protein with global MS severity outcome (i.e., positive correlation coefficient of the CSF protein was translated as transcription level correlating positively with pathway activation). Next, we calculated average gene expression and %-age of cells expressing the gene for all genes in pyroptosis pathway. The linear regression model *(log_2_(1+y) ∼ x: x = cell frequency expressing gene, y = average gene expression of gene)* was constructed to identify outliers (> 2SD or < −2SD of residual) that were excluded from further analysis. Finally, we calculated tPPAS of each cell type in MS and control tissue using following formula:

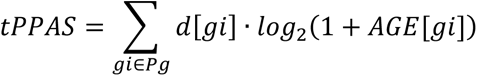

AGE: average gene expression; d: directionality; Pg: genes in pathway

## Data Availability

All data produced in the present study are available upon reasonable request to the authors.

## Acknowledgements

We thank all our patients, their caregivers, and our clinical team at the Neuroimmunological Diseases Section without whom this work would not be possible. We also thank the Rocky Mountain Laboratories Genomics Unit of the NIAID Research Technologies Branch for kind assistance with RNA-seq.

## Author contributions

Conceptualization: BB; Methodology: JK, PK, SA, RG-M, KP, BB; Investigation: JK, PK, SA, BB; Visualization: JK, PK, SA, BB; Funding acquisition: BB; Critical input to data interpretation: BB, SMH, RG-M, KP; Supervision: BB, KP, RG-M, VF; Resources: NP, VF; Writing original draft: JK, PK, SA, BB; Review & scientific editing: all authors

## Data and materials availability

All RNA-seq data that support the findings of this study will be made publicly available through NCBI’s Gene Expression Omnibus (GEO) (accession#) upon publication.

## Supplementary Materials

Material and methods-In-vitro mechanistic studies

Figs. S1 to S9

Table S1

## Other Supplementary Material for this manuscript includes the following

Table S2 Table S3

