## Supplemental Information for "Clemastine fumarate accelerates accumulation of disability in progressive multiple sclerosis by enhancing pyroptosis"

**One Sentence Summary:** Clemastine enhanced disability accumulation in patients progressing by non-lesional MS activity by potentiating intrathecal P2RX7 signaling and pyroptosis.

**Supplementary Materials**

**Material and methods**

**In-vitro mechanistic studies**

**Reagents**

The reagents/growth factors used in the study are as follows: adenosine 5′-triphosphate disodium salt hydrate, ATP (Millipore Sigma, A6419), biotin (Sigma-Aldrich, 4639), clemastine fumarate salt, CLM (Sigma-Aldrich, SML0445), insulin solution, human (Sigma-Aldrich, 19278), JNJ-54175446 (MedChemExpress, HY-117508), lipopolysaccharides from Escherichia coli O111:B4, LPS (Sigma-Aldrich, L4391), neurotrophin 3 (NT3; EMD Millipore, GF031), nigericin sodium salt (Sigma-Aldrich, N7143), recombinant human IGF-I (R&D Systems, 291-G1-200), recombinant human HGF (R&D Systems, 294-HG-025), recombinant human macrophage colony stimulating factor, M-CSF (PeproTech, 300-25), recombinant human platelet derived growth factor, PDGF-AA (R&D Systems, 221-AA-050), 3,3,5-Triiodo-l-thyronine, T3 (Sigma-Aldrich, T2877).

**Cell cultures**

THP-1 and THP1-KO-GSDMD cells were purchased from InvivoGen (#thp-null and #thp-kogsdmdz, respectively; CA, USA). The cells were grown in 1640 RPMI medium supplemented with 2 mM L-glutamine, 25 mM HEPES, 10% FBS (v/v), penicillin (100 U/mL) and streptomycin (100 µg/mL), and 100 µg/mL Normocin at 37°C with 5% $CO_2$. The THP1-KO-GSDMD cells were maintained in a growth medium supplemented with the selective antibiotic 100 µg/mL Zeocin™ following every other passage. Experiments were performed on the cells with less than 10 passages.

Monocytes elutriated from human peripheral blood were obtained from the National Institutes of Health Blood Bank. Monocytes were cultured in RPMI 1640 medium supplemented with 10% FBS (v/v), 1% GlutaMax, penicillin (100 U/mL), and streptomycin (100 µg/mL). For differentiation to macrophages (MDMs), the cells were cultured for 7 days in the presence of recombinant human M-CSF (50 ng/mL).

Human iPSC-derived oligodendrocytes were generated using previously published protocol (*29*) Neurospheres enriched for OLIG2 glia progenitors were plated into 96-well plate coated with poly-L-ornithine and laminin (one sphere per well) and cells were allowed to migrate out of the sphere. The cells were differentiated in “PDGF medium” containing morphogens that specifically promote oligodendrocyte differentiation and maturation (DMEM/F12 medium containing non-essential amino acids (1x), GlutaMAX (1x), 2-mercaptoethanol, penicillin (100 U/mL) and streptomycin (100 µg/mL), supplemented with N2 supplement (1×), B27 supplement (1×), 10 ng/mL PDGFaa, 10 ng/mL IGF-1, 5 ng/mL HGF, 10 ng/mL NT3, 60 ng/mL T3, 100 ng/mL Biotin, 1 µM cAMP, 25 µg/mL insulin) for the next 40 days. Two-thirds of the medium was changed every other day.

**IL-1β release**

Quantification of secreted IL-1β was performed using Human IL-1 beta/IL-1F2 Quantikine ELISA Kit (R&D Systems, DLB50) according to the kit instructions.THP-1 and THP1-KO-

GSDMD cell culture supernatants were diluted 2-fold, while MDM cell culture supernatants were 100-fold diluted. The samples were analyzed at 450 nm with a reference reading at 540 nm with the use of Infinite ® 200 PRO microplate reader.

**Caspase-1 activity**

After drug treatment, the culture medium was collected, centrifuged at 250 x *g* for 5 min to remove debris, and used immediately. Caspase-1 activity in cell culture supernatants was assayed using Caspase-Glo 1 Inflammasome Bioluminescent Assay (Promega, G9952) per the manufacturer's recommendations. Assay specificity was conferred by inclusion of a proteasome inhibitor MG-132 in the lytic reagent and using a caspase-1 inhibitor Ac-YVAD-CHO at concentration of 5 µM and 25 µM. Luminescence was recorded using Promega Microplate Reader GloMax® Explorer GM3500.

**Cell membrane permeability**

THP-1 cells were plated in ultra-low attachment, U-bottom plates, primed with 200 ng/mL of LPS overnight, and then stimulated either with medium + DMSO (Ctrl), 10 µg/mL Clemastine (CLM) or 2 mM ATP +/- 10 µg/mL CLM in the presence of 12.5 nM SytoxGreen (Invitrogen™, S7020) for 90 min. The SYTOX® Green–stained cells were subjected to fluorescence analysis using 488 nm excitation on flow cytometry Aurora 3 Laser V/B/R – 38 Channel System. MDM cells were plated in 35-mm glass bottom dishes, primed with 200 ng/mL of LPS overnight, and then stimulated either with medium + DMSO (Ctrl), 10 µg/mL Clemastine (CLM), 2 mM ATP +/- 10 µg/mL CLM or 10 µM Nigericin for 6h. Sytox Green (1µM) was added to the medium for monitoring cell membrane integrity. After 15 min, images of pyroptotic cells were captured using a Leica DMIL LED Fluorescence Microscope equipped with green 488 nm laser at room temperature. The pictures were processed by the ZEN 2012 Image program.

**Cell death assay**

Lytic cell death was assayed by using CytoTox 96 Non-Radioactive Cytotoxicity Assay (Promega, G1780) according to the manufacturer's protocol. The absorbance at 492 nm was measured as an indicator of LDH activity in cell culture supernatants with the use of Infinite ® 200 PRO microplate reader. The percentage of cytotoxicity was calculated with the following formula: Percent Cytotoxicity = 100 × (Experimental Sample − Culture Medium Background) / (Maximum LDH release − Culture Medium Background).

**Cell viability**

The cell viability was determined by CellTiter 96® AQueous One Solution Cell Proliferation Assay (Promega, G3580) as per the manufacturer's instruction. Following treatment incubation, the medium was removed and replaced with 100 µL of fresh medium containing 20 µL of MTS reagent. After 2 h incubation with MTS, the absorbance was measured at 490 nm using Infinite ® 200 PRO microplate reader, and the percentage of viable cells was calculated using the following formula: Percent Viability = 100 × (Experimental Sample) / (Control sample).

**Bulk RNA-seq**

*Sample preparation and RNA extraction:* For bulk RNA-seq, the cells were plated in 6-well plates ($1x10^6$ per well), primed with 200 ng/mL of LPS overnight, and then stimulated either with medium + DMSO (Ctrl) or 2 mM ATP +/- 10 µg/mL CLM for 3 h or 6h. For each drug condition we sequenced the following samples (n=3): 1) MDMs incubated with drugs for 3h; 2) MDMs incubated with drugs for 6h and 3) THP-1 cells incubated with drugs for 3 h. Total RNA extraction was performed using Trizol LS reagent (Invitrogen). Three volumes of Trizol LS reagent were added to each cell pellet ($1x10^6$), lysate was combined with 200 µL of 1-Bromo-3-chloropropane (MilliporeSigma, St. Louis, MO), and RNA aqueous phase was obtained according to the manufacturer's recommendations (ThermoFishcer Scientific). RNA containing aqueous phase was combined with 600 µL of RLT lysis buffer (Qiagen, Valencia, CA) with 1% beta mercaptoethanol (MilliporeSigma, St. Louis, MO) and RNA was extracted using Qiagen AllPrep DNA/RNA 96-well system (Valencia, CA). An additional on-column DNAse 1 treatment was performed during RNA extraction. The RNA integrity was verified with Agilent RNA 6000 Pico kit on the 2100 Bioanalyzer (Agilent Technologies) according to the manufacturer's protocol. RNA was quantitated using a fluorescence assay (Quant-it RiboGreen RNA, Thermofisher Scientific, Waltham, MA) on a Tecan Spark multiplate reader (Tecan, Switzerland).

*Library preparation and RNA sequencing:* The Stranded mRNA Prep kit (Illumina) was used to generate sequencing libraries according to the manufacture's reference guide #1000000124518, v02. Briefly, 10 ng (sample 9-MDMs_ATP+CLM_6hr) or 25 ng (all others) total RNA was used as template for library preparation, using 16 PCR cycles to enrich for adapter-modified products. Final libraries were pooled in equimolar concentrations and sequenced as paired-end 2 X 74 bp reads on two NextSeq 550 instrument runs using the High Output 150 cycle sequencing kit (Illumina). Sequencing was performed on 2 NextSeq 550® (Illumina) 74 cycle + 74 cycle symmetrical runs.

### Figures

**Primary monocyte - derived macrophages**

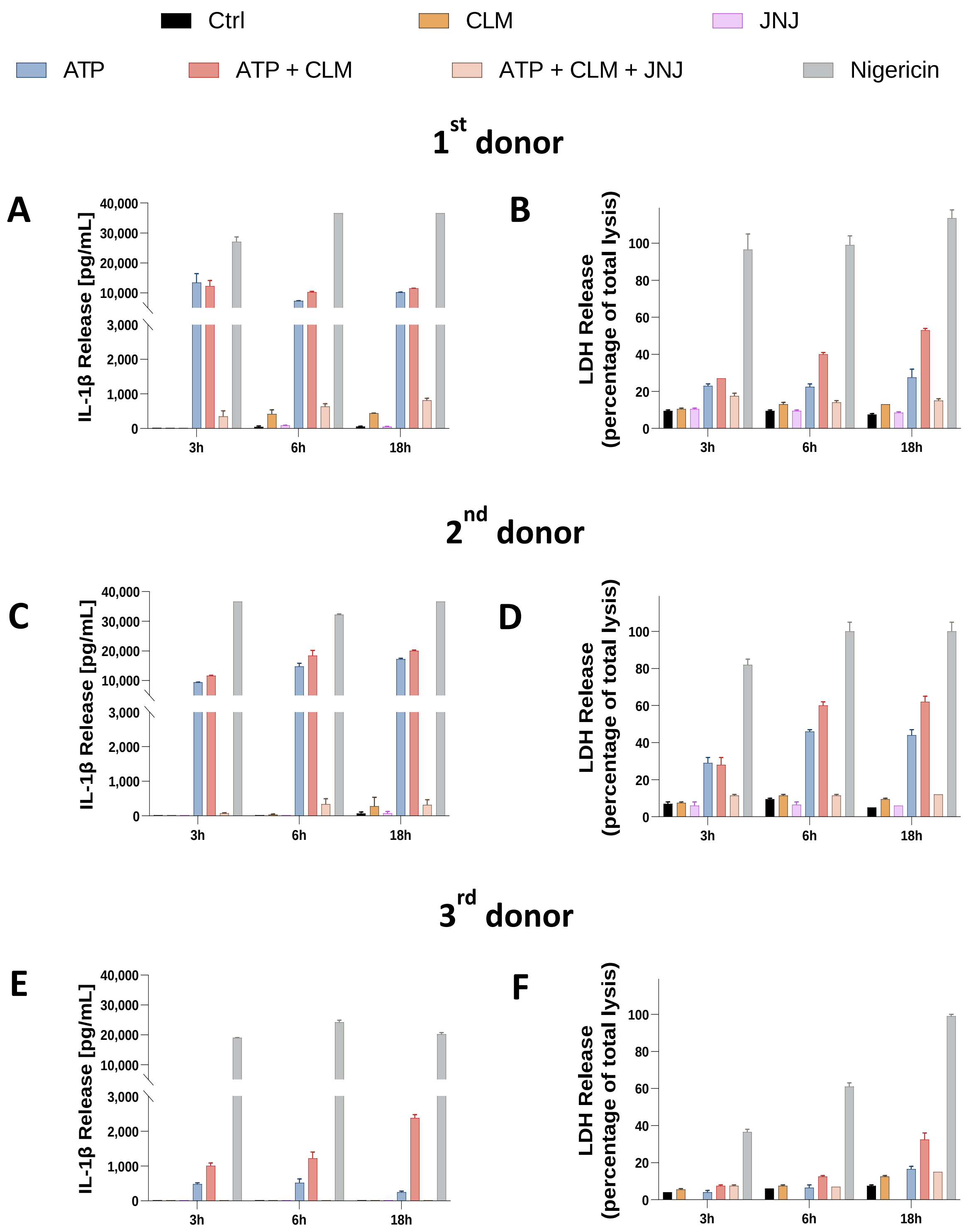

**Fig. S1. The effect of Clemastine (CLM) on ATP-induced pro-inflammatory cytokine IL-1β release and lytic cell death of primary monocyte-derived macrophages (MDMs) presented for each donor individually.** The MDM cells were primed with 200 ng/mL LPS overnight, then pre-treated with 30 nM JNJ-54175446 (a selective purine P2X7 receptor antagonist) for 1 h, followed by treatment either with medium + DMSO (negative control, Ctrl), 10 µg/mL CLM, 2 mM ATP +/- 10 µg/mL CLM or 10 µM Nigericin (positive control) for 3h, 6h, and 18 h. Levels of pro-inflammatory cytokine IL-1β **(A,C,E**) and LDH activity **(B,D,F**) in the culture supernatants. Data are presented as mean ± SEM of independent experiment performed in duplicate.

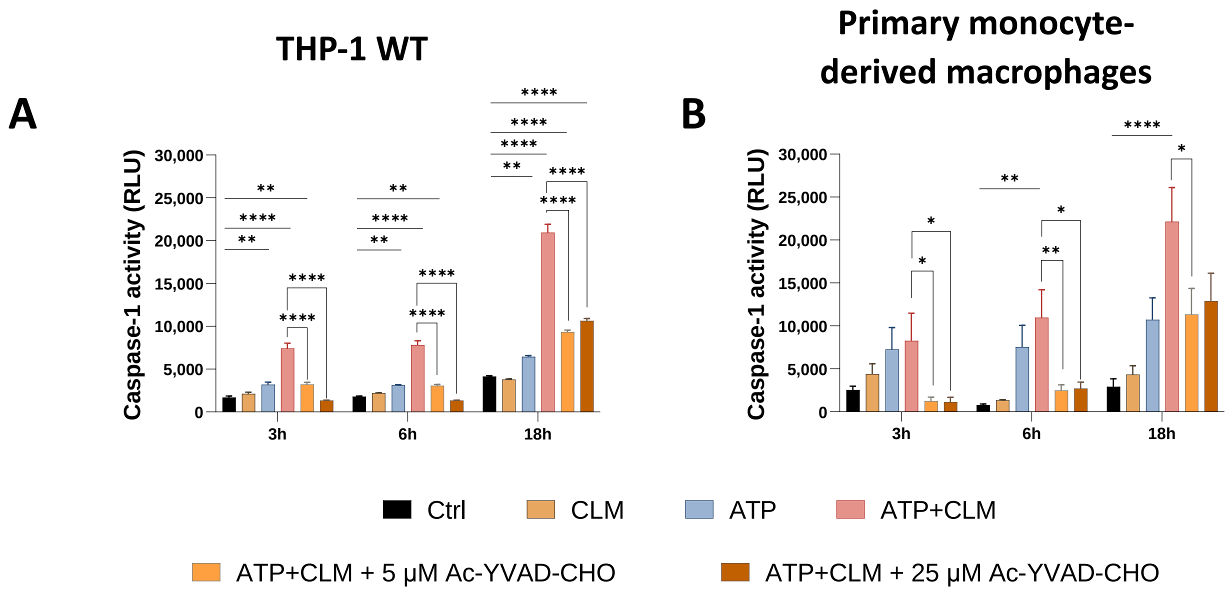


**Fig. S2. Caspase-1 Assay Specificity.** The cells were primed with 200 ng/mL LPS overnight followed by treatment either with medium + DMSO (negative control, Ctrl), 10 µg/mL CLM, 2 mM ATP, 2 mM ATP+10 µg/mL CLM with/without Ac-YVAD-CHO (5 µM and 25 µM) for 3h, 6h, and 18 h. Where indicated, the Ac-YVAD-CHO inhibitor was added to the lytic Caspase-Glo® 1 reagent to confirm the specificity of the luminescent caspase-1 signal. Activity of Caspase-1 was evaluated by bioluminescence assay in the cell culture supernatants of THP-1 cells (A) and primary human monocyte-derived macrophages (B). Data are presented as mean ± SEM of three independent experiments performed in duplicate. One way ANOVA followed by Dunnett's multiple comparisons test was used to compare the testing groups with control group (Ctrl). One-way ANOVA test followed by Holm-Sidak's multiple comparison was used to compare ATP+CLM and ATP+CLM with Ac-YVAD-CHO inhibitor group (to assess the Caspase-1 assay specificity). *, $P \leq 0.05$. **, $P \leq 0.01$. ***, $P \leq 0.001$. ****, $P \leq 0.0001$.

**MYC Mediated Apoptosis Signaling**

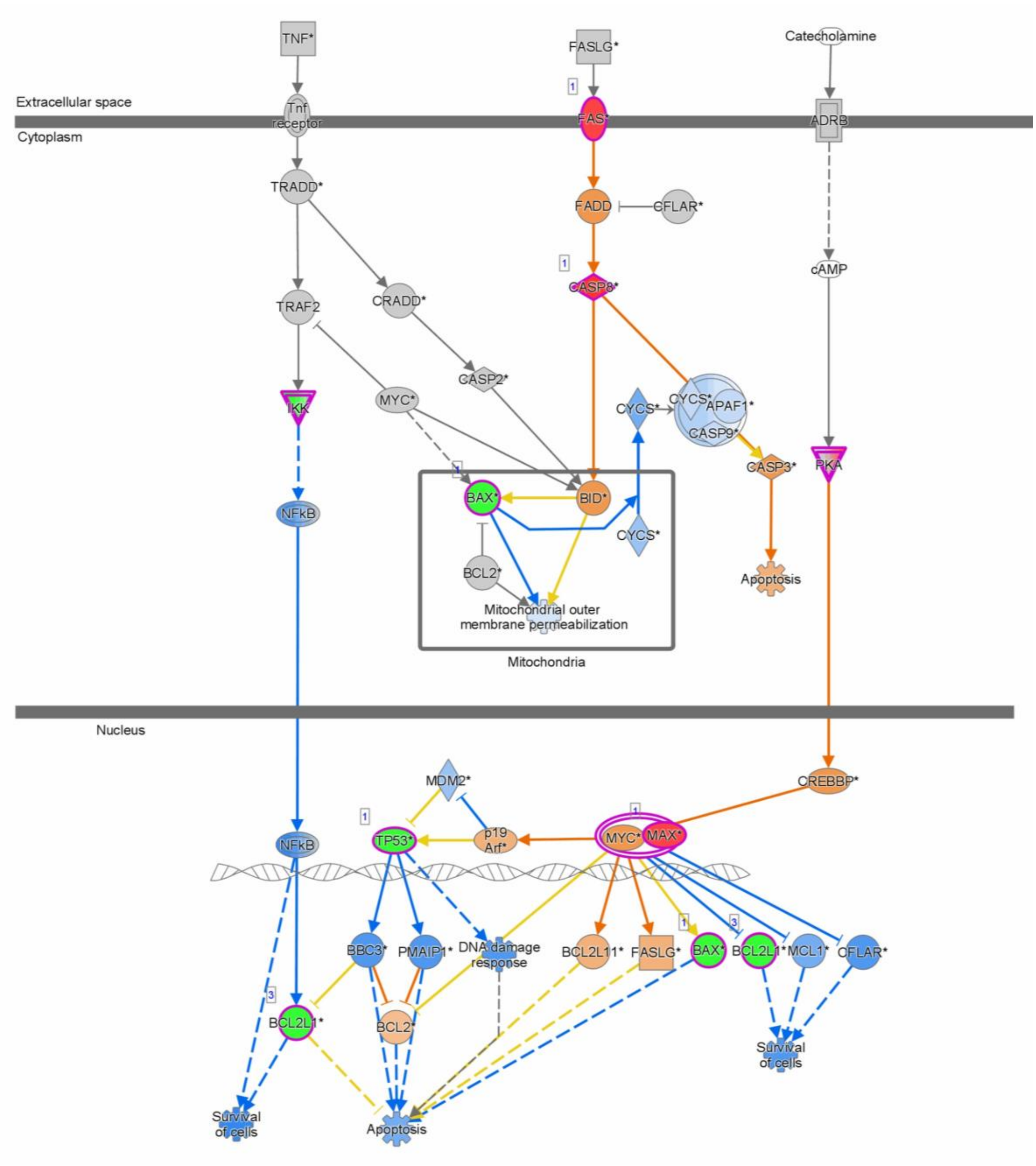


**Fig. S3. Mitochondrial dysfunction and apoptosis pathways activated by ATP treatment predicted by ingenuity pathway analysis.** mRNAs with FDR < 0.05 and log2 fold change > 0.6 was included in the activated pathways.

**Inflammasome pathway**

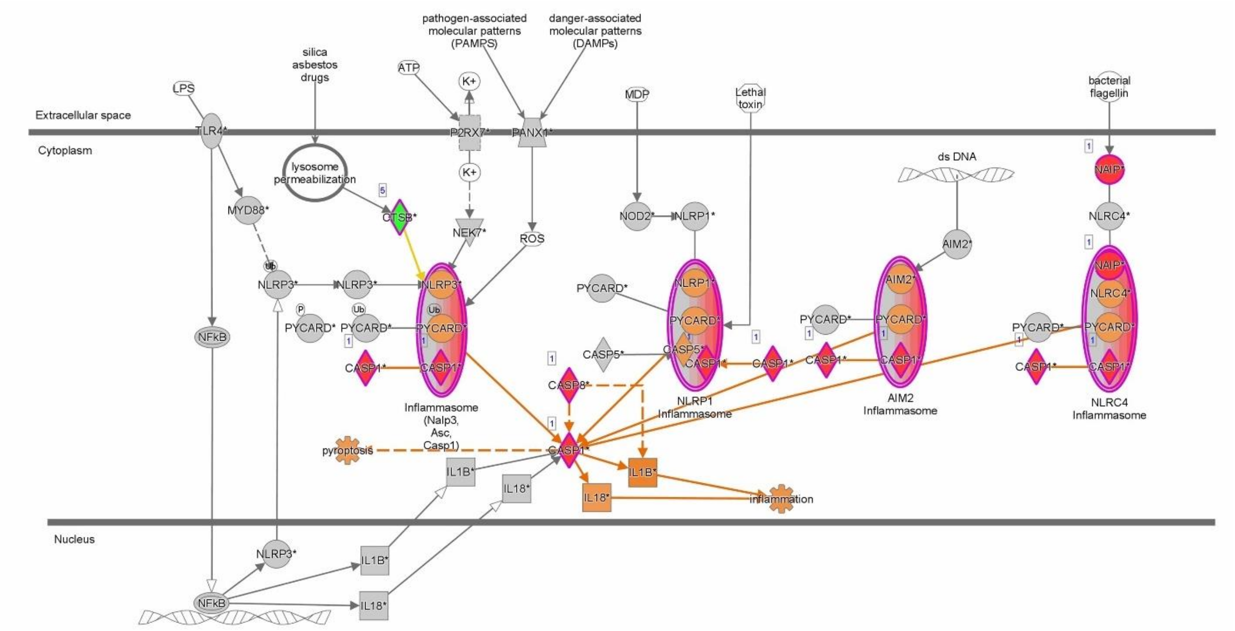


**Fig. S4. The inflammasome signaling pathway activated by clemastine+ATP treatment predicted by ingenuity pathway analysis.** mRNAs with FDR < 0.05 and log2 fold change > 0.6 was included in the activated pathways.

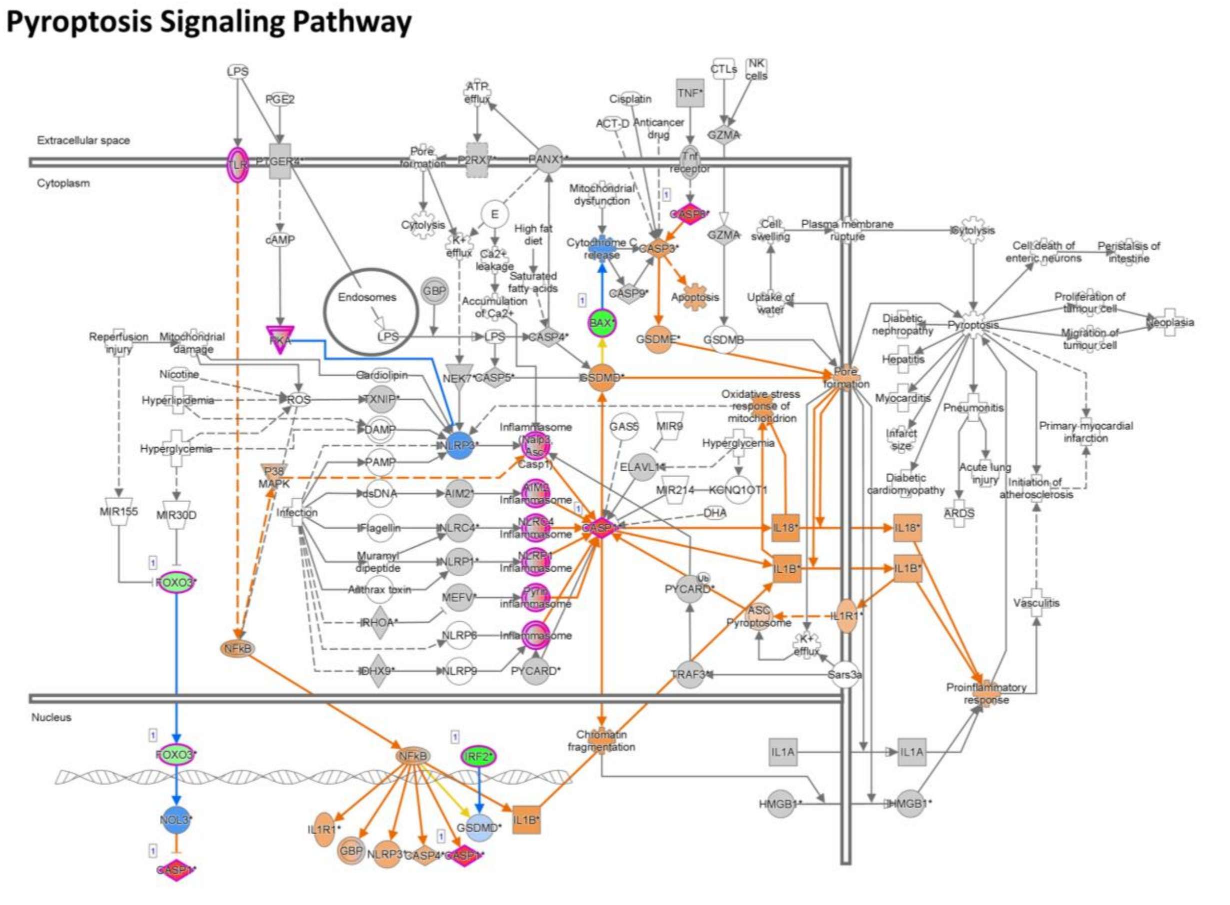


**Fig. S5. The pyroptosis signaling pathway activated by clemastine+ATP treatment predicted by ingenuity pathway analysis**. mRNAs with FDR < 0.05 and log2 fold change > 0.6 was included in the activated pathways.

### Immunogenic Cell Death Signaling Pathway

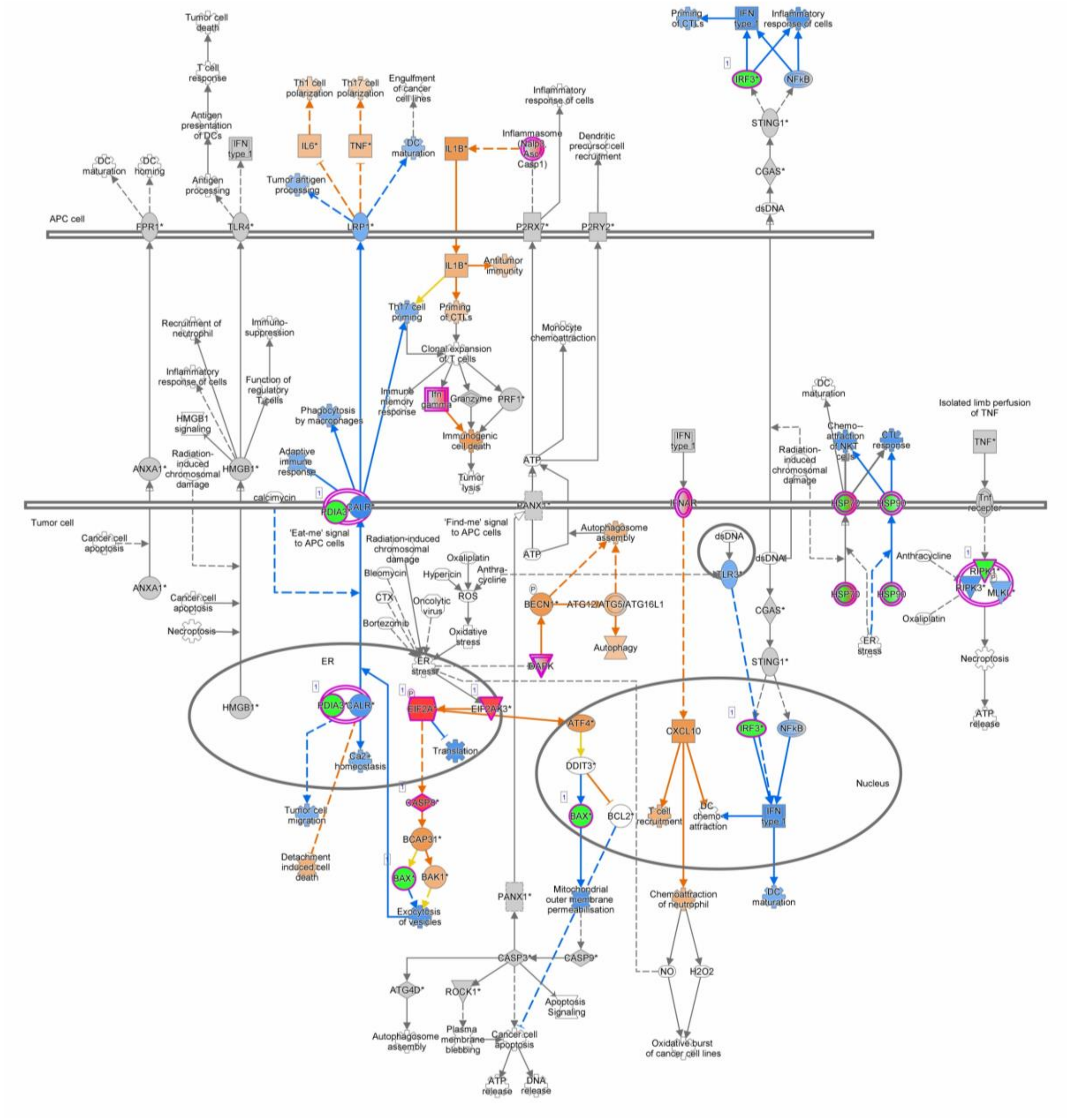

**Fig. S6. The immunogenic cell death pathway activated by clemastine+ATP treatment predicted by ingenuity pathway analysis.** mRNAs with FDR < 0.05 and log2 fold change > 0.6 was included in the activated pathways.

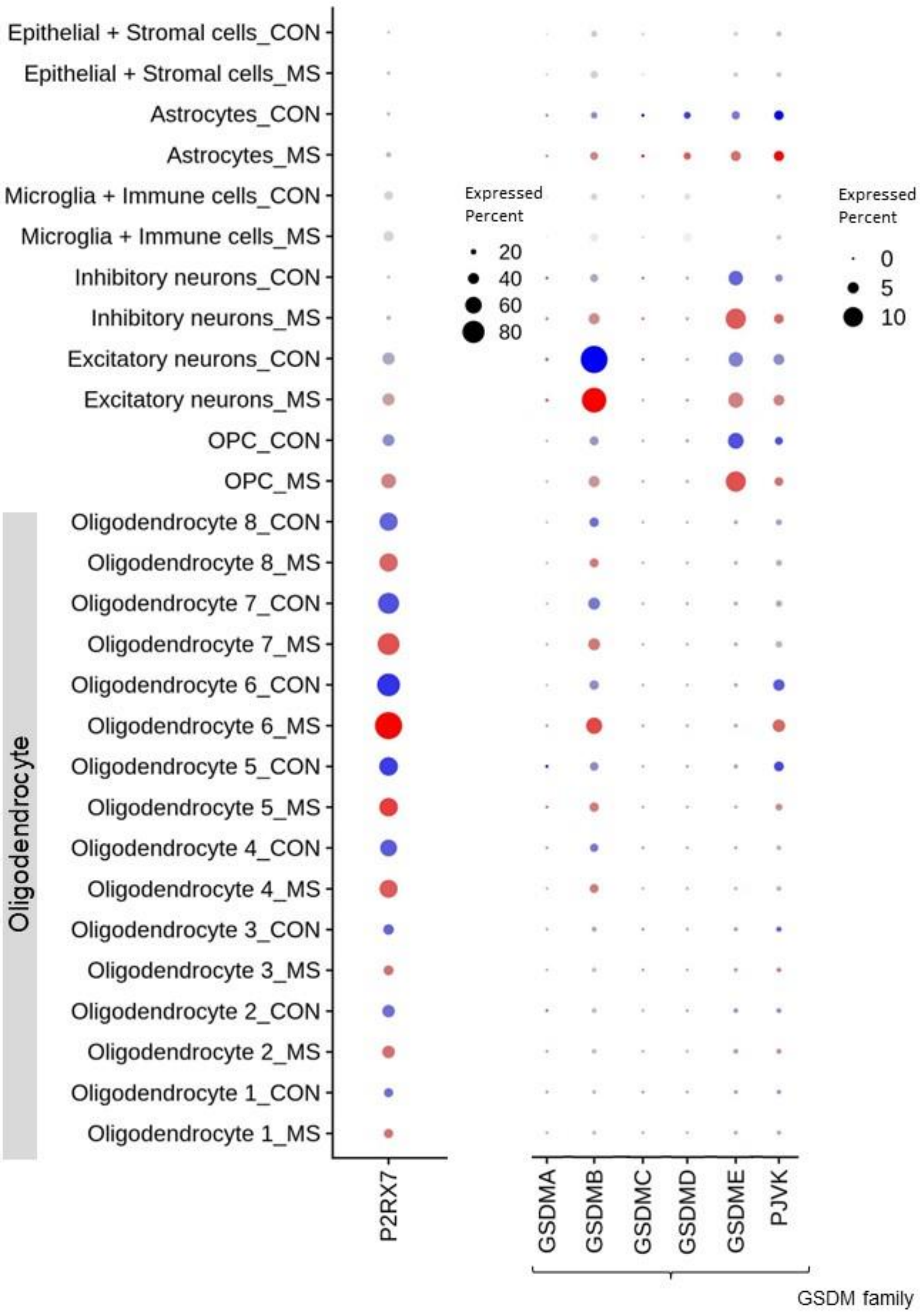


**Fig. S7. Gene expression associated with pyroptosis signaling pathway of CNS cell types: MS vs. control tissue**. Figure shows gene expressions based on cell types from MS (red dots) and control tissue (blue dots). P2RX7: purinergic receptor P2X7; GSDM: gasdermin; PJVK: pejvakin; NAWM: normal-appearing white matter; OPC: oligodendrocyte precursor cells.

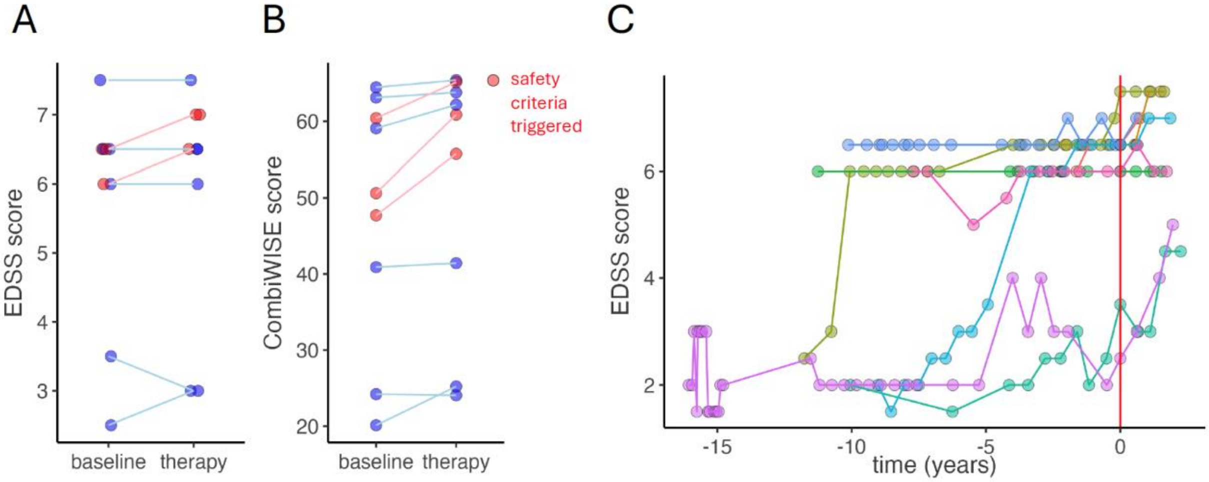


**Fig. S8. Measurement of clinical disability in MS patients receiving clemastine treatment.** Levels of clinical disability of nine patients treated with clemastine with at least one follow-up visit 6 months after therapy initiation were assessed by EDSS (**A**) and CombiWISE (**B**) at baseline (before clemastine initiation) and on therapy (6 month after clemastine start). Patients that triggered safety criteria are labeled in red. (**C**) Natural history EDSS data collected over >15 years; start of clemastine therapy is depicted with red vertical line.

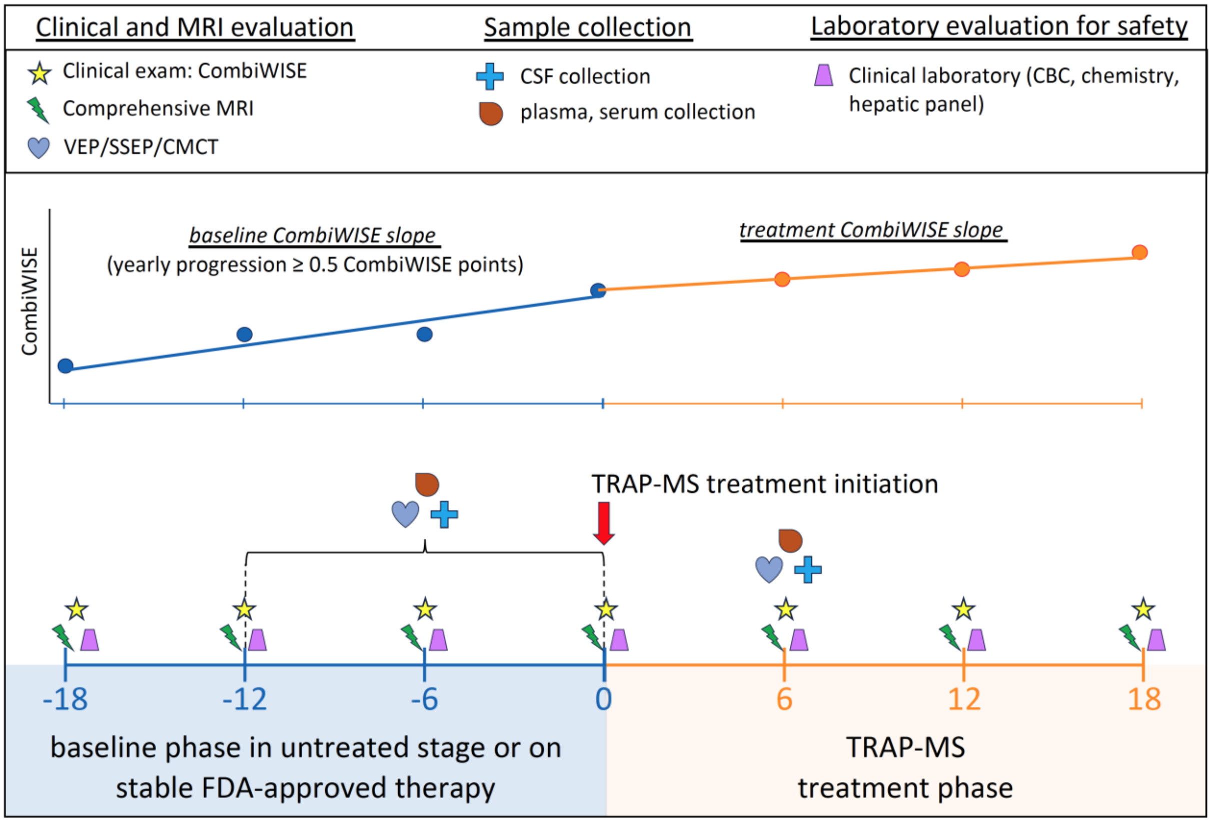


**Fig. S9. Design of TRAP-MS trial.** Patients were screened under the Natural History protocol for minimum of 18 months with minimum of 4 visits separated by at least 6 months on stable FD-approved therapy or untreated. At each visit, clinical exam, comprehensive MRI, and a panel of clinical laboratory tests were performed. Patients with calculated yearly CombiWISE change greater than qualified for TRAP-MS trial. Abnormal results from Visual Evoked Potential (VEP), Somatosensory Evoked Potential (SSEP) or Central Motor Conduction Time (CMCT) during baseline were required to start Clemastine as a TRAP-MS therapy. Baseline lumbar puncture (associated with plasma and serum collection) was performed within 12 months of TRAP-MS therapy initiation. After therapy initiation patients were followed every six months with clinical exam, MRI, and laboratory tests. Six months after therapy initiation an on-therapy lumbar puncture was performed (with associated plasma and serum collection) as well as electrophysiological test that was abnormal during the baseline. CombiWISE values on therapy were used to calculate therapy CombiWISE slope.

Table S1: Demographic data

| ***Cohort*** | | TRAP-MS clemastine | TRAP-MS other therapy | HD | Natural History cross-sectional | Placebo longitudinal |
|---|---|---|---|---|---|---|
| ***patients (N)*** | | 9 | 42 | 49 | 168 | 72 |
| ***diagnosis (%)*** | | | | | | |
| | RR-MS | 22.2% | 14.3% | NA | 29.8% | 4.2% |
| | PP-MS | 33.3% | 52.4% | NA | 51.8% | 15.3% |
| | SP-MS | 44.4% | 33.3% | NA | 18.5% | 80.6% |
| ***sex (%)*** | | | | | | |
| | F | 55.6% | 50.0% | 46.9% | 58.3% | 52.8% |
| | M | 44.4% | 50.0% | 53.1% | 41.7% | 47.2% |
| ***Age (years)*** | | | | | | |
| | mean | 62.5 | 58.9 | 38.5 | 53.2 | 57.5 |
| | SD | 12.1 | 8.5 | 13.5 | 12.1 | 7.3 |
| | range (min-max) | 35.1-73.3 | 36.2-76.5 | 19.4-71.3 | 19.5-74.9 | 38.4-74.6 |
| ***Disease duration (years)*** | | | | | | |
| | mean | 22.3 | 18.5 | NA | 13.9 | 15.3 |
| | SD | 11.8 | 12.3 | NA | 10.9 | 10.4 |
| | range (min-max) | 8.1-48.2 | 5.3-48.1 | NA | 0.1-54.1 | 0.1-42.2 |
| ***EDSS*** | | | | | | |
| | mean | 5.7 | 5.4 | 0.9 | 5.0 | 5.3 |
| | SD | 1.6 | 1.4 | 0.9 | 1.7 | 1.6 |
| | range (min-max) | 2.5-7.5 | 2.0-7.0 | 0.0-3.0 | 1.5-8.0 | 1.5-8.0 |
